# Dependence-aware aggregation maps shared genetic architecture across cardiovascular traits

**DOI:** 10.64898/2026.11.24.26368884

**Authors:** Richard M. Monaghan, Simon G. Williams, Hui Guo, Bernard D. Keavney

## Abstract

**Background:** Shared genetic effects are pervasive across clinically related cardiovascular diseases and can expose biology that crosses diagnostic boundaries, but correlation between related phenotypes (shared patients, risk factors, overlapping definitions) makes naive cross-trait recurrence hard to interpret because evidence is counted repeatedly. We asked which genes carry coordinated positive association across a complete cardiovascular panel beyond the recurrence expected from trait dependence.

**Methods:** We developed the Cross-Trait Aggregation Score (CTAS), a per-gene statistic applied to MAGMA gene-level association across 16 Pan-UK Biobank cardiovascular traits. Panel dependence was estimated as a transformed rank-correlation matrix; each gene’s 16-trait profile was decorrelated by ZCA-cor whitening to remove the expected correlation; and CTAS combined the magnitude and effective breadth of positive residual evidence across the whitened coordinates. The statistic was calibrated against an empirical null of 10^9^ simulated profiles, with genome-wide Benjamini–Hochberg false-discovery-rate control, after which significant genes were merged into loci and characterised by clinical-family composition, expression colocalisation, comparison with prior multi-trait studies and FinnGen.

**Results:** CTAS prioritised 223 genes at 91 loci (q<0.05), with nested sets of 99 genes at 46 loci (q<0.01) and 38 genes at 21 loci (q<0.001); membership was identical across independent null simulations. The loci formed an overlapping multi-family continuum rather than discrete disease classes: 88% spanned at least two clinical families and the fraction spanning four or more increased from 21% to 43% across the nested sets. Expression colocalisation supported 31 of 91 loci (31 of 87 evaluable). Literature convergence with four prior multi-trait studies covered 58 of 91 loci and 17 of the 21 most stringent loci. In FinnGen, the proportion of CTAS genes with external support across at least three of six clinical families of cardiovascular endpoints increased across the nested sets, from 49% at q<0.05 to 57% at q<0.01 and 61% at q<0.001, against 6% of background genes.

**Conclusions:** CTAS evaluates a complete correlated trait panel in one calibrated test per gene, discounting the recurrence expected from trait correlation. In cardiovascular disease, it prioritised genes and loci with multi-family, independently supported evidence, providing a focused starting point for mechanistic and translational follow-up.

## Introduction

Shared genetic effects across multiple traits (pleiotropy) are a pervasive feature of complex disease, not an exception [1, 2]. Across related diseases, they can mark points of biological convergence, help explain patterns of comorbidity, connect apparently distinct clinical outcomes and reveal biology obscured when phenotypes are analysed separately [3–5]. The challenge is to recover this evidence without mistaking expected correlation for shared biology.

Cardiovascular disease provides a compelling setting in which to recover this shared architecture [6, 7]. Coronary, cerebrovascular, myocardial, valvular, hypertensive and thrombotic phenotypes share developmental programmes, vascular and cardiac biology and inflammatory pathways [8, 9]. Their comorbidity [10, 11], overlapping diagnostic definitions, shared participants and common analytical structure also create substantial cross-trait dependence. Consequently, association signals across cardiovascular traits are not independent [4, 12]. This creates both the scientific opportunity and the analytical difficulty: treating the traits independently can count the same underlying evidence repeatedly, whereas pruning related traits removes clinically meaningful structure. An analysis therefore needs to keep the complete panel while separating coordinated evidence from the recurrence expected under dependence.

Existing cross-trait methods address complementary parts of this problem [13, 14]. Multi-Trait Analysis of GWAS summary statistics (MTAG) uses genetic correlation to increase power for trait-specific discovery [15]. Association analysis based on subsets (ASSET) identifies the subset of traits supporting a variant association [16]. Pleiotropic analysis under the composite null hypothesis (PLACO) tests pairwise pleiotropic association [17]. Colocalisation methods such as HyPrColoc assess whether regional signals are compatible with a shared causal variant [18]. Genomic SEM models latent genetic structure across phenotypes [19]. Gene- and SNP-set methods have also extended multivariate association testing to gene-level units [20–22]. PLEIO performs joint SNP-level mapping across correlated traits while accounting for genetic correlation, heritability and covariance structure [23]. These approaches have distinct inferential targets and outputs (Table 1). The Cross-Trait Aggregation Score (CTAS), introduced here, complements them by asking a different question: once panel dependence has been discounted, how much positive gene-level evidence remains, and how widely is it spread? It answers with one calibrated score per gene across all tested genes.

**Table 1.** Conceptual comparison of CTAS with established multi-trait genetic methods. Methods are compared by unit of analysis, question addressed, treatment of cross-trait structure, primary output and inferential niche. The comparison is descriptive rather than a ranking; the methods address complementary questions. Abbreviations and methodological details are given in the table footnote.

| Method | Unit of analysis | Question addressed | Role of cross-trait structure | Primary output | Inferential niche |
|---|---|---|---|---|---|
| MTAG | SNP | Which variants associate with a focal trait when discovery is augmented by cross-trait genetic correlation? | Genetic correlations reweight and combine per-trait summary statistics, increasing effective sample size for per-trait association | Trait-specific SNP association statistics with boosted power | Boosting single-trait GWAS discovery using correlated traits |
| ASSET | SNP | Which subset of traits or studies supports a shared association at a given variant? | Heterogeneity across traits is modelled; the best-fitting trait subset is selected per variant | Subset-based meta-analytic association statistics per SNP, with the contributing trait subset identified | Identifying the trait subset driving a shared variant signal |
| PLACO | SNP | Does a variant show pleiotropic association across two traits, after accounting for their correlation? | Trait correlation calibrates the null distribution for the product of per-trait Z-scores | Per-SNP pleiotropic association P-value | Pairwise pleiotropic signal detection at the variant level |
| HyPrColoc | Genomic region | Do multiple traits share a single causal variant at a given locus? | LD structure and per-trait GWAS summary statistics are used to estimate the posterior probability of colocalization within a genomic window | Per-locus posterior probability of colocalization; likely shared causal variant identified | Resolving whether locus-level signals share a causal variant |
| Genomic SEM | SNP / latent structure | What latent factor structure underlies the genetic architecture of a trait family, and how do variants load on it? | A structural equation model is fitted to the genetic covariance matrix to decompose it into common and trait-specific factors | Latent factor loadings and factor-specific GWAS statistics | Decomposing shared genetic structure into interpretable latent dimensions |
| Multi-trait gene/SNP-set tests (MTaSPUsSet, MSATS, OMGA) | Gene or SNP set | Is a gene or variant set jointly associated with multiple correlated traits? | Incorporates trait dependence and, where applicable, LD while aggregating evidence across traits and variants | Omnibus multi-trait gene/SNP-set association P value | Joint detection of gene/set association across multiple phenotypes |
| PLEIO | SNP / locus | Is a variant jointly associated with multiple traits after accounting for trait architecture? | Models genetic correlation, heritability and environmental/error covariance | Joint PLEIO association P value and locus-level association profile | Joint mapping and interpretation of pleiotropic loci |
| CTAS | Gene profile across the complete panel | Which genes retain the strongest positive cross-trait evidence after expected panel dependence is discounted? | Trait correlation matrix Σ = 2 sin(πρS/6); ZCA-cor whitening of each gene's one-sided trait z-scores; only positive whitened evidence is retained | Gene-level aggregate evidence (CTAS) across the 16-trait panel; empirical P and genome-wide BH q | Ranked dependence-aware gene architecture; no causal-variant, common-effect or causal-gene claim |

SNP, single-nucleotide polymorphism; LD, linkage disequilibrium; GWAS, genome-wide association study; SEM, structural equation modelling; CTAS, Cross-Trait Aggregation Score; Σ, 16 × 16 trait correlation matrix, 2 sin(πρS/6) transform of the Spearman rank correlations ρS of the sixteen trait signals; ZCA-cor, zero-phase component analysis whitening of the correlation matrix (Σ^−1/2); P, empirical P value; BH, Benjamini–Hochberg. CTAS is the method introduced in this study. CTAS identifies aggregate cross-trait gene-level evidence; it does not itself establish shared causal variants or estimate a common effect size.

CTAS evaluates each gene once across the prespecified full panel, without searching for phenotype subsets. Panel dependence is estimated as a transformed rank-correlation matrix, and each gene profile is decorrelated by ZCA-cor whitening, so recurrence predictable from trait correlation is discounted before scoring; the whitened coordinates are mixtures of traits, not named phenotypes. CTAS then combines the magnitude and effective breadth of the remaining positive evidence and is calibrated against an empirical null of 10^9^ simulated profiles before false-discovery-rate control. Biological context is evaluated separately, through clinical-family profiles, colocalisation and external cardiovascular support.

Here, we applied CTAS to gene-level association statistics for 16 cardiovascular traits from the Pan-UK Biobank [24] (Table 2). We asked whether the complete cardiovascular panel could resolve a shared gene- and locus-level signal beyond that expected from trait dependence. CTAS prioritised 223 genes mapping to 91 loci at q<0.05, with nested sets of 99 genes at 46 loci at q<0.01 and 38 genes at 21 loci at q<0.001. Across simulation, sensitivity, cardiovascular-family, colocalisation and external-support analyses, we evaluated the calibration, robustness and biological informativeness of these discoveries. The prioritised loci did not separate into disease-specific classes. Instead, they overlapped across clinical families, their family coverage broadened with CTAS stringency, and they carried colocalisation, literature and FinnGen support.

**Table 2.** The prespecified 16-trait cardiovascular panel. The sixteen both-sex cardiovascular phenotypes that constitute the fixed CTAS analysis panel, listed in canonical order with the abbreviations used throughout the paper. For each trait, the Pan-UK Biobank phenotype identifier (ICD-10 or phecode) indexes the exact summary-statistic source, alongside all-ancestry case counts, European-ancestry case and control counts, and SNP-based heritability on the observed and liability scales. This panel and its canonical abbreviations define the traits that populate the eight cardiovascular trait-family groupings used in the subsequent analyses. Abbreviations and scale definitions are given in the table footnote.

| **Trait** | **Pan-UKBB phenotype** | **Sample size** |  |  | **SNP heritability, EUR** |  |
|---|---|---|---|---|---|---|
|  |  | **Cases, all ancestries** | **EUR cases** | **EUR controls** | **Observed scale, h<sup>2</sup> (SE)** | **Liability scale, h<sup>2</sup> (SE)** |
| Coronary artery disease (CAD) | ICD-10 I25 | 32,926 | 31,148 | 389,383 | 0.0362 (0.0032) | 0.1263 (0.0112) |
| Myocardial infarction (MI) | ICD-10 I21 | 10,595 | 10,141 | 410,390 | 0.0147 (0.0026) | 0.1076 (0.019) |
| Angina pectoris (Ang) | ICD-10 I20 | 22,491 | 21,286 | 399,245 | 0.0306 (0.0028) | 0.1356 (0.0124) |
| Atrial fibrillation (AF) | ICD-10 I48 | 19,554 | 19,218 | 401,313 | 0.0298 (0.0034) | 0.1411 (0.0161) |
| Heart failure (HF) | ICD-10 I50 | 7,800 | 7,448 | 413,083 | 0.0042 (0.0021) | 0.0383 (0.0192) |
| Cardiomyopathy (CMP) | ICD-10 I42 | 1,662 | 1,662 | 418,869 | 0.0015 (0.002) | 0.043 (0.0573) |
| Hypertension (HTN) | Phecode 401 | 99,977 | 94,311 | 325,488 | 0.0862 (0.0049) | 0.1672 (0.0095) |
| Atherosclerosis (Ath) | ICD-10 I70 | 933 | 933 | 419,598 | 0 (0.0021) | -0.0016 (0.0958) |
| Other peripheral vascular disease (I73) | ICD-10 I73 | 4,930 | 4,778 | 415,753 | 0.0085 (0.0026) | 0.1076 (0.0329) |
| Ischaemic stroke (IS) | ICD-10 I63 | 4,437 | 4,275 | 416,256 | 0.0015 (0.0021) | 0.0206 (0.0289) |
| Transient ischaemic attack (TIA) | ICD-10 G45 | 3,208 | 3,150 | 417,381 | 0.0024 (0.0023) | 0.0417 (0.04) |
| Phlebitis/ thrombophlebitis (I80) | ICD-10 I80 | 4,886 | 4,746 | 415,785 | 0.0091 (0.003) | 0.1158 (0.0382) |
| Venous embolism/ thrombosis (I82) | ICD-10 I82 | 514 | 514 | 420,017 | 0.0011 (0.0021) | 0.082 (0.1565) |
| Aortic aneurysm (AoAn) | ICD-10 I71 | 2,031 | 2,031 | 418,500 | 0.0009 (0.0022) | 0.022 (0.0538) |
| Aortic valve disease (AoV) | ICD-10 I35 | 3,552 | 3,479 | 417,052 | 0.0083 (0.0022) | 0.1336 (0.0354) |
| Mitral valve disease (MV) | ICD-10 I34 | 3,216 | 3,153 | 417,378 | 0.0019 (0.0024) | 0.033 (0.0417) |

EUR, European ancestry; h<sup>2</sup>, SNP-based heritability; SE, standard error. Observed-scale heritability reflects raw trait variance captured by common SNPs; liability-scale heritability adjusts for disease prevalence. Phenotype codes identify the exact Pan-UK Biobank GWAS summary statistic used. I80 and I82 denote the complete three-character Pan-UK Biobank ICD-10 categories (phlebitis and thrombophlebitis; other venous embolism and thrombosis), including all subcodes, and are distinct from the broader FinnGen venous thromboembolism endpoint (I9\_VTE) used for external support.

## Methods

### Study design and cardiovascular trait panel

We conducted a secondary analysis of publicly available genome-wide association study (GWAS) summary statistics from the Pan-UK Biobank meta-analysis outputs [24] for a prespecified panel of 16 cardiovascular traits (Table 2). The panel retained each phenotype as a separate analytical input so that shared gene-level association evidence could be evaluated while accounting for dependence between related traits. No individual-level participant data were accessed.

The primary analysis was performed on a matrix of gene-level association P values for 19,080 genes across the 16 traits. We developed CTAS as a single dependence-aware statistic applied to each full-panel gene profile, without searching across phenotype subsets. Genes were ranked using empirically calibrated CTAS P values and grouped into nested false-discovery-rate tiers before locus assignment. The resulting locked gene and locus sets were then used for simulation and sensitivity analyses, cardiovascular-family characterisation, colocalisation and evaluation against previous studies and FinnGen; none of these downstream analyses contributed to the initial selection of genes or loci.

The analysis proceeded in four stages. First, trait-specific GWAS summary statistics were converted into a common gene-level association matrix. Second, CTAS was defined, calibrated and used to prioritise genes and loci across the complete cardiovascular panel. Third, the locked CTAS gene and locus sets were tested for robustness and family-level structure. Fourth, the same locked sets were evaluated against expression colocalisation, previous multi-trait cardiovascular studies and FinnGen.

### GWAS processing and gene-level association analysis

Summary statistics from the Pan-UK Biobank multi-ancestry meta-analysis field were obtained in February 2026 for the 16 both-sex phenotypes listed in Table 2. Pan-UK Biobank reports meta-analysis association P values as neglog10_pval_meta; these values were converted to P values on the original scale before gene analysis. Variant identifiers formatted as chromosome, position, reference allele and alternate allele were mapped to rsIDs using the European panel of the 1000 Genomes Project phase 3 reference BIM file [25]. Variants that could not be matched were excluded before gene analysis.

Gene-level association P values were calculated separately for each trait using MAGMA v1.10 [26] and the SNP-wise mean model. Genes were defined using the NCBI build 37.3 annotation on GRCh37/hg19, with variants assigned within the gene body and no additional flanking window; this restricted assignment to variants within the annotated gene interval and limited overlap-based inclusion of neighbouring genes. Linkage disequilibrium was estimated using the European 1000 Genomes reference panel [25]. Thus, the discovery association P values came from the Pan-UK Biobank meta-analysis field, whereas linkage-disequilibrium correction used a European reference panel. A common sample-size parameter of 500,000 was specified for the primary MAGMA analyses so that the same gene-level analysis definition was applied across all traits. The effect of replacing this common value with phenotype-specific effective sample sizes was examined separately as a sensitivity analysis.

The resulting trait-specific outputs were combined by gene symbol to form a 19,080-gene by 16-trait matrix. P values were required to be finite and within the interval [0,1]. Missing gene–trait values were retained when estimating cross-trait dependence using all available pairwise observations and were subsequently treated as providing no evidence in the gene-level CTAS calculation. In total, 3,144 of 305,280 gene–trait cells were missing, and 777 genes lacked a value for at least one trait. Matrix completeness and trait-pair overlap were recorded as quality-control measures (Supplementary Table 12).

### Estimation of cross-trait dependence

Dependence between traits was estimated from the genome-wide gene-level P-value matrix using a rank-based Gaussian working model [27]. Pairwise Spearman correlations ρ_S_ between traits were calculated across genes from the gene-level P-value profiles using pairwise-complete observations, with ties assigned their average rank. Each was converted to the Gaussian scale to give the 16 × 16 transformed rank-correlation matrix *R* = 2sin(*πρ*_S_/6) (applied elementwise, with unit diagonal) [28]. This correlation matrix parameterised the multivariate normal working null used for simulation and whitening. Its minimum eigenvalue was 0.50, so R was positive definite and the prespecified projection to the nearest positive-definite correlation matrix [29] was not required.

For whitening, we used ZCA-cor whitening, that is, the symmetric inverse square root of the correlation matrix. Writing its eigendecomposition as *R* = *VΛV*^T^, where V is the matrix of eigenvectors and Λ is the diagonal matrix of eigenvalues λ_j_, the whitening matrix is

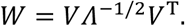

Because the minimum eigenvalue of R was 0.50, W was computed directly from this eigendecomposition. An eigenvalue floor (10^−10^) retained in the code as a numerical safeguard never activated, and the floored and unfloored whitening matrices were identical. W removes the correlation expected under the fitted gene-level null while avoiding an order-dependent decomposition. ZCA-cor whitening maximises the correlation between each standardised original variable and its whitened counterpart [30] and retains the original coordinate ordering, allowing its coordinates to be used as trait-aligned residual dimensions for descriptive summaries. Nevertheless, each coordinate remains a linear combination of the full panel and should not be interpreted as an independent named phenotype.

### CTAS statistic

For each gene, MAGMA gene-level P values were clipped to the interval [10^−300^, 1 − 10^−10^] and converted to upper-tail normal deviates,

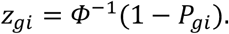

Missing values were assigned *z*_g*i*_ = 0, corresponding to no positive association evidence. The gene vector was then whitened as **z̃**_g_ = *W***z**_g_. Positive truncation was applied after whitening. Because the input P values are one-sided, a negative whitened coordinate represents less evidence on that axis than predicted from the gene’s evidence on correlated traits, whereas a positive coordinate represents excess evidence; CTAS retains only this excess positive evidence. A half-normal or other truncating transformation applied before decorrelation would remove this distinction and would not prevent negative whitened coordinates. For each whitened coordinate, positive evidence was therefore represented by

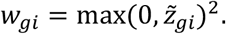

The magnitude component was defined as

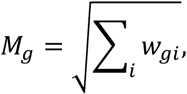

and effective breadth was calculated using the participation ratio [31],

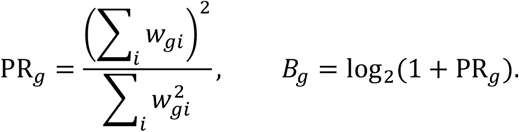

CTAS was the product

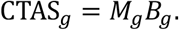

CTAS was set to zero when no whitened coordinate was positive. The magnitude component summarises the overall amount of positive residual evidence, whereas the breadth component describes how that evidence is distributed across decorrelated dimensions. Because whitening mixes information across correlated traits, neither individual whitened coordinates nor negative whitened residuals were assigned a direct biological interpretation.

### Empirical calibration and false-discovery-rate control

The primary CTAS null distribution was generated by drawing 10^9^ vectors from *N*(**0**, *R*) using a fixed random seed of 42; the complete analysis was repeated with seeds 1 and 2. Each simulated vector was subjected to the same ZCA-cor whitening and CTAS calculation as an observed gene. For an observed value *c*_g_, the directly resolved empirical P value was

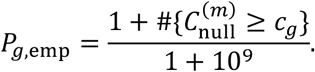

This plus-one correction prevented zero empirical P values and gave a direct resolution limit of 1/(10^9^ + 1) [32]. Three genes exceeded every null draw. For the most extreme of these, LPA, an exponential model fitted to the upper tail of the simulated null survival function gave an estimated P value below this limit. The other two retained the floor value, and all remaining genes retained directly counted empirical P values. Tail handling affected no q-value threshold: the empirical P values at the three set boundaries (5.8×10^−4^, 5.1×10^−5^ and 2.0×10^−6^) lay far above the resolution limit (Supplementary Fig. 1; Supplementary Table 2).

Empirical or extrapolated P values, as applicable, were adjusted across all 19,080 genes using the Benjamini–Hochberg procedure [33]. We defined nested FDR-controlled gene sets at q<0.05, q<0.01 and q<0.001. These thresholds provided progressively stringent discovery tiers used throughout the analyses (Supplementary Table 1). Gene and locus membership at all three thresholds was identical for null seeds 42, 1 and 2 (Supplementary Table 2).

Throughout the Results and Discussion, loci refer to frozen genomic neighbourhoods constructed from significant CTAS genes using the prespecified positional merging rule described below. Locus representative refers to the highest-CTAS significant gene within that neighbourhood, not to an inferred causal gene.

### Locus construction and representative genes

Significant genes were assigned to a genomic-neighbourhood map constructed before the final CTAS analysis from a fixed reference-gene set and 101 prespecified reference intervals. A reference gene was assigned to an existing interval when its NCBI build 37.3 coordinates lay within 1 Mb of that interval. Reference genes not assigned to any of the 101 intervals were ordered by chromosome and grouped de novo when successive gene intervals were separated by no more than 1 Mb, using the running maximum end coordinate. Non-significant reference genes were retained in the map and could bridge adjacent regions. Final significant genes were looked up in this fixed map and counted as CTAS hits only when they passed the relevant q-value threshold. Regions were sorted deterministically by chromosome, start coordinate, end coordinate and representative gene and assigned fixed sequential identifiers, which are retained for the 91 loci containing q<0.05 genes.

Within each locus, the statistical representative was the significant gene with the highest CTAS; in every locus this was also the gene with the smallest q value, and no ties occurred. The locus map and representative definitions were frozen before downstream characterisation. Representatives were used to provide one reproducible index gene per region and were not interpreted as causal-gene assignments. Locus membership and the nested locus tiers are reported in Supplementary Table 3.

### Null calibration and statistic-behaviour simulations

Simulation analyses were conducted in whitened space, where null vectors follow a 16-dimensional standard normal distribution under the fitted model. Calibration was assessed using 200,000 independently generated null vectors (seed 20260805), evaluated against a separate 1,000,000-vector reference null. Because these simulations are specified in whitened space, where the null is N(0, I_16_) whatever correlation matrix is whitened, they characterise the behaviour of the statistic; the primary analysis itself used the 10^9^-draw null. Observed rejection proportions were compared with nominal probabilities of 0.2, 0.1, 0.05, 0.02, 0.01, 0.005 and 0.001 (Supplementary Table 4).

We next tested how CTAS behaved when the same total signal was distributed over different numbers of decorrelated axes. For total non-centrality *λ* ∈ {20,40,80} and active-axis count *k* = 1, …, 16, the mean on each active axis was set to 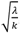, with the remaining axes centred at zero. We generated 30,000 replicates per condition (seed 77), calculated CTAS from the same draws, and normalised mean CTAS to its value at *k* = 1. A complementary simulation crossed per-axis means of 0.5, 1, 1.5, 2 and 3 with active-axis counts of 1, 2, 4, 8, 12 and 16, using 20,000 replicates per cell (seed 131). Together, these simulations characterised the statistical calibration and intended response of CTAS to signal magnitude and spread; biological organisation of the prioritised loci was evaluated separately using clinical-family, colocalisation and external-support analyses (Fig. 2; Supplementary Fig. 2; Supplementary Table 4).

**Figure 1.**
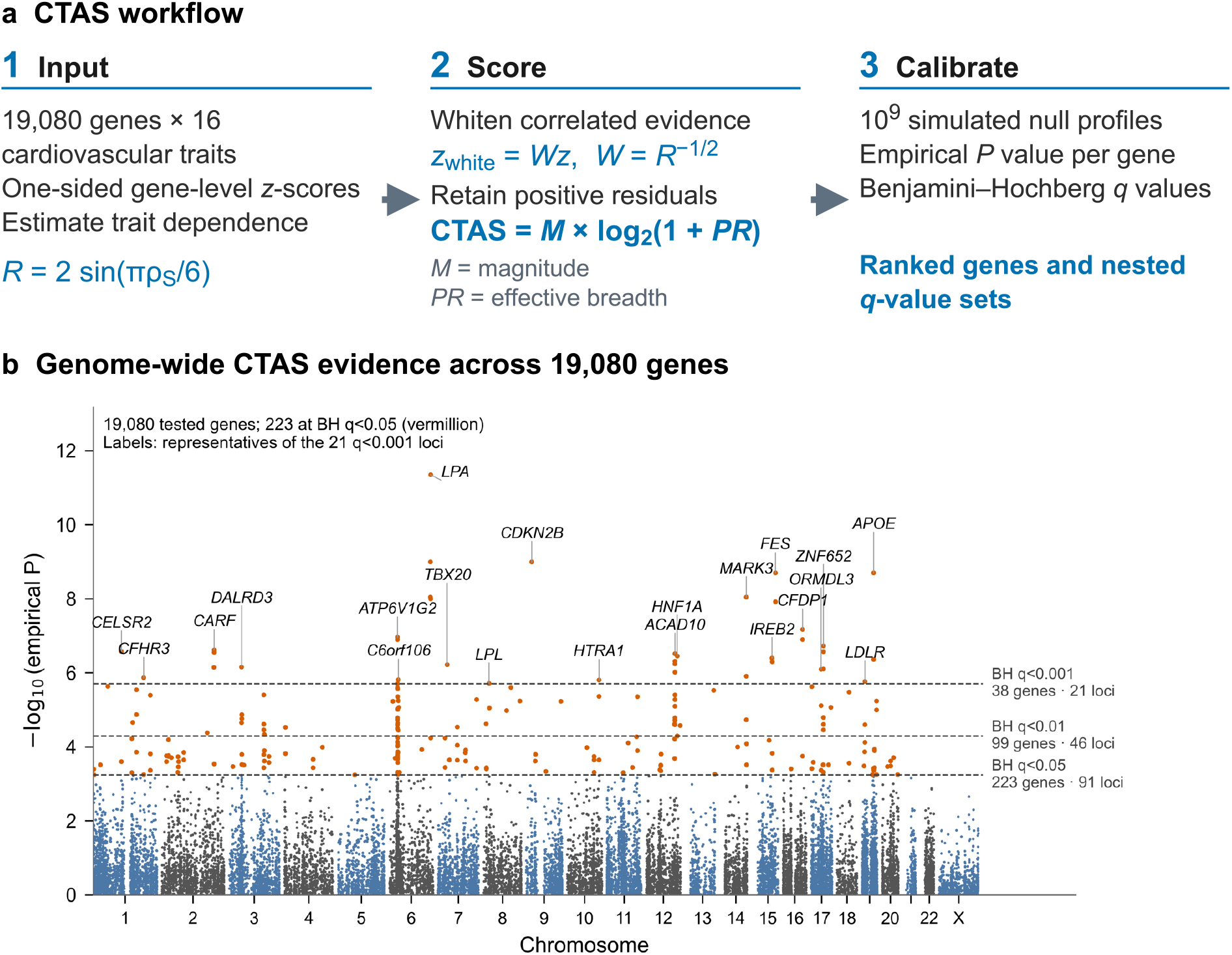
CTAS method and genome-wide results. (a) CTAS comprises three stages. First, the input is a matrix of one-sided gene-level z-scores for 19,080 genes across 16 cardiovascular traits, with panel dependence represented by the transformed rank-correlation matrix R = 2 sin(πρS/6). Second, ZCA-cor whitening adjusts each gene profile for expected dependence as z_white = Wz, where W = R^(−1/2). Only positive residual evidence is retained. CTAS combines its magnitude M and effective breadth PR as CTAS = M × log2(1 + PR); whitened axes are mathematical mixtures of traits, not phenotypes. Full definitions are given in the Methods. Third, the observed CTAS is compared with 10⁹ simulated null profiles processed identically to obtain an empirical P value, followed by genome-wide Benjamini–Hochberg q across all 19,080 genes. (b) Genome-wide CTAS evidence across 19,080 genes. The 223 genes with BH q<0.05 are vermillion. Dashed lines mark the largest empirical P value within each nested BH-q set (q<0.05, q<0.01 and q<0.001), comprising 223, 99 and 38 genes at 91, 46 and 21 loci. Labels identify the statistical representatives of the 21 q<0.001 loci; representatives are locus labels, not causal-gene assignments. Membership was identical across null seeds 42, 1 and 2.

**Figure 2.**
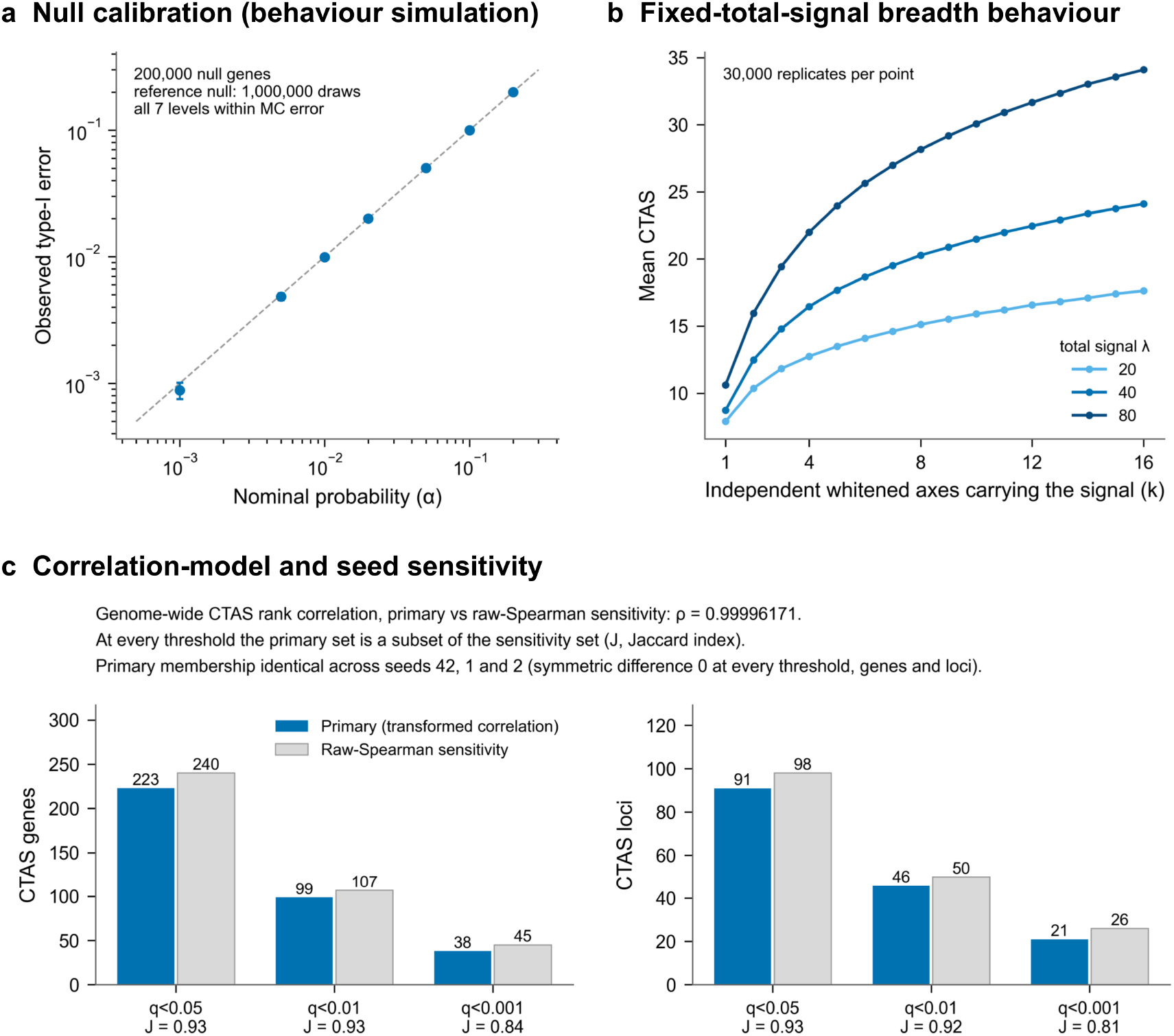
Calibration, intended behaviour and sensitivity of CTAS. (a) Null calibration (method-behaviour diagnostic): observed versus nominal type-I error for 200,000 independent null genes evaluated against a 1,000,000-draw reference null; all seven levels fall within Monte Carlo error (bars, 95% intervals). This simulation characterises the statistic; the primary analysis itself uses the 10⁹-draw null. (b) Fixed-total-signal breadth behaviour: with total non-centrality λ held fixed, spreading the signal across more independent whitened axes (k) increases mean CTAS (30,000 replicates per point). This shows the intended breadth sensitivity of the statistic; it does not by itself demonstrate biological pleiotropy. Simulations are specified in whitened space, where the null is N(0, I16) whatever trait correlation matrix is whitened. (c) Correlation-model and seed sensitivity. Primary analysis (transformed correlation matrix): 223 / 99 / 38 genes and 91 / 46 / 21 loci at q<0.05 / q<0.01 / q<0.001. Raw-Spearman sensitivity: 240 / 107 / 45 genes and 98 / 50 / 26 loci; at every threshold the primary set is a subset of the sensitivity set (Jaccard J shown). The genome-wide CTAS rank correlation between the two analyses is ρ = 0.99996171. Primary gene and locus membership is identical across null seeds 42, 1 and 2 (symmetric difference 0).

### Correlation-model and seed sensitivity

As a sensitivity analysis, the complete primary analysis was repeated with the raw Spearman matrix in place of the transformed rank-correlation matrix, using the same ZCA-cor whitening, 10^9^-draw null, seeds and Benjamini–Hochberg procedure. Agreement was assessed by genome-wide Spearman rank correlation of CTAS and by set overlap at each threshold. Gene and locus membership of the primary analysis was compared across null seeds 42, 1 and 2 (Fig. 2c; Supplementary Tables 2 and 5).

### Effective-sample-size sensitivity analysis

To assess dependence on the common sample-size parameter used in the primary MAGMA analyses, we repeated the complete gene-level analysis using a phenotype-specific effective sample size,

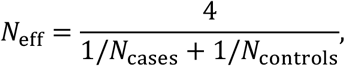

for each case–control trait, using the total case and control counts across the ancestries contributing to the corresponding Pan-UK Biobank meta-analysis. Variant inputs, genome build, gene annotation, linkage-disequilibrium reference and SNP-wise mean model were unchanged. The resulting 19,080 × 16 P-value matrix and the common-sample-size matrix were scored with the same sensitivity pipeline (raw Spearman working correlation matrix, ZCA-cor whitening, 10^6^-draw null, Benjamini–Hochberg correction and the locked locus map). Gene-rank correlation and changes in the status of the final CTAS genes and loci at each q-value threshold were recorded (Supplementary Table 6).

### Trait-composition sensitivity analyses

We performed 16 leave-one-trait-out analyses. In each analysis, one trait was removed, a new 15 × 15 rank-based working dependence matrix and symmetric whitening transform were estimated, and a new 1,000,000-vector null distribution was simulated with seed 42. CTAS P values were recalculated and Benjamini–Hochberg correction was reapplied across all 19,080 genes. Upper-tail extrapolation was not required for the retention boundaries in these analyses. A locked locus was considered retained at a given q-value threshold if at least one of its corresponding full-analysis member genes passed that threshold after trait omission; retention of the canonical representative itself was recorded as a secondary measure (Supplementary Table 7). The leave-one-trait-out, effective-sample-size and reduced-panel analyses used their recorded raw Spearman working correlation matrices and 10^6^-draw nulls and were not rerun under the primary 10^9^-draw analysis; their results were evaluated against the primary gene and locus sets.

We also performed a reduced-panel analysis after removing six overlapping coronary, peripheral vascular and cerebrovascular manifestations: myocardial infarction, angina, other peripheral vascular disease (ICD-10 I73), atherosclerosis, ischaemic stroke and transient ischaemic attack. This retained coronary artery disease as the single representative of the broader atherosclerotic and obstructive disease cluster. The other retained traits were atrial fibrillation, aortic aneurysm, aortic valve disease, cardiomyopathy, heart failure, hypertension, mitral valve disease, phlebitis/thrombophlebitis (ICD-10 I80) and venous embolism/thrombosis (ICD-10 I82). A new 10×10 dependence matrix, whitening transform and 1,000,000-vector null calibration were fitted, followed by the same exponential-tail and multiple-testing procedures used in the primary analysis. Primary regional retention required at least one reduced-panel significant gene to map to a frozen genomic neighbourhood from the full analysis; retention of a locked member gene was assessed secondarily. Regions detected only in the reduced panel were retained as diagnostics and were not added to the canonical locus set (Supplementary Table 8).

### Cardiovascular-family architecture

For descriptive interpretation, the 16 traits were grouped a priori into eight cardiovascular families: coronary disease (coronary artery disease, myocardial infarction and angina), pressure (hypertension), arrhythmia (atrial fibrillation), heart failure (heart failure and cardiomyopathy), valve disease (aortic and mitral valve disease), aortic and peripheral vascular disease (aortic aneurysm, other peripheral vascular disease [ICD-10 I73] and atherosclerosis), cerebrovascular disease (ischaemic stroke and transient ischaemic attack), and thrombosis (phlebitis/thrombophlebitis, ICD-10 I80, and venous embolism/thrombosis, ICD-10 I82).

We calculated two distinct family summaries for each canonical locus representative. First, descriptive family coverage was determined from the original MAGMA results: a family was counted when at least one of its component traits had P<0.01 for the representative gene. This measure was not used to select genes or loci. Second, continuous family composition was calculated from the positive components of the representative’s ZCA-cor-whitened vector. For trait-aligned residual coordinate *t*, *v_t_* = max(0, *z̃_t_*). Unsquared residuals were used for this descriptive composition, in contrast to the squared weights used to calculate CTAS. The contribution of family *f* was the mean positive evidence across its *n_f_* component traits,

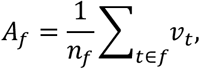

and its share was *S_f_* = *A_f_*/ Σ*_f_A_f_*. The effective number of represented families was defined as 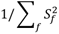. This family-share diversity measure is distinct from the participation-ratio breadth component of CTAS. Root-mean-square aggregation within families was assessed as a sensitivity analysis. These profiles provide a size-adjusted descriptive summary of the ZCA-cor-whitened coordinates and were not treated as a causal decomposition of trait effects.

For ordination, family shares were Hellinger-transformed [34] as 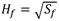, centred without scaling and analysed by principal-components analysis. We evaluated whether locus profiles formed discrete classes using Ward’s hierarchical clustering, k-means clustering with 50 random starts and partitioning around medoids for *k* = 2, …, 10. Candidate solutions were assessed using mean silhouette width [35] and agreement between algorithms using the adjusted Rand index [36]. Stability of partitioning around medoids was additionally assessed over 200 bootstrap iterations, each sampling 80% of loci. Clustering of the untransformed family shares was examined as a sensitivity analysis (Fig. 3; Supplementary Fig. 3; Supplementary Table 3).

**Figure 3.**
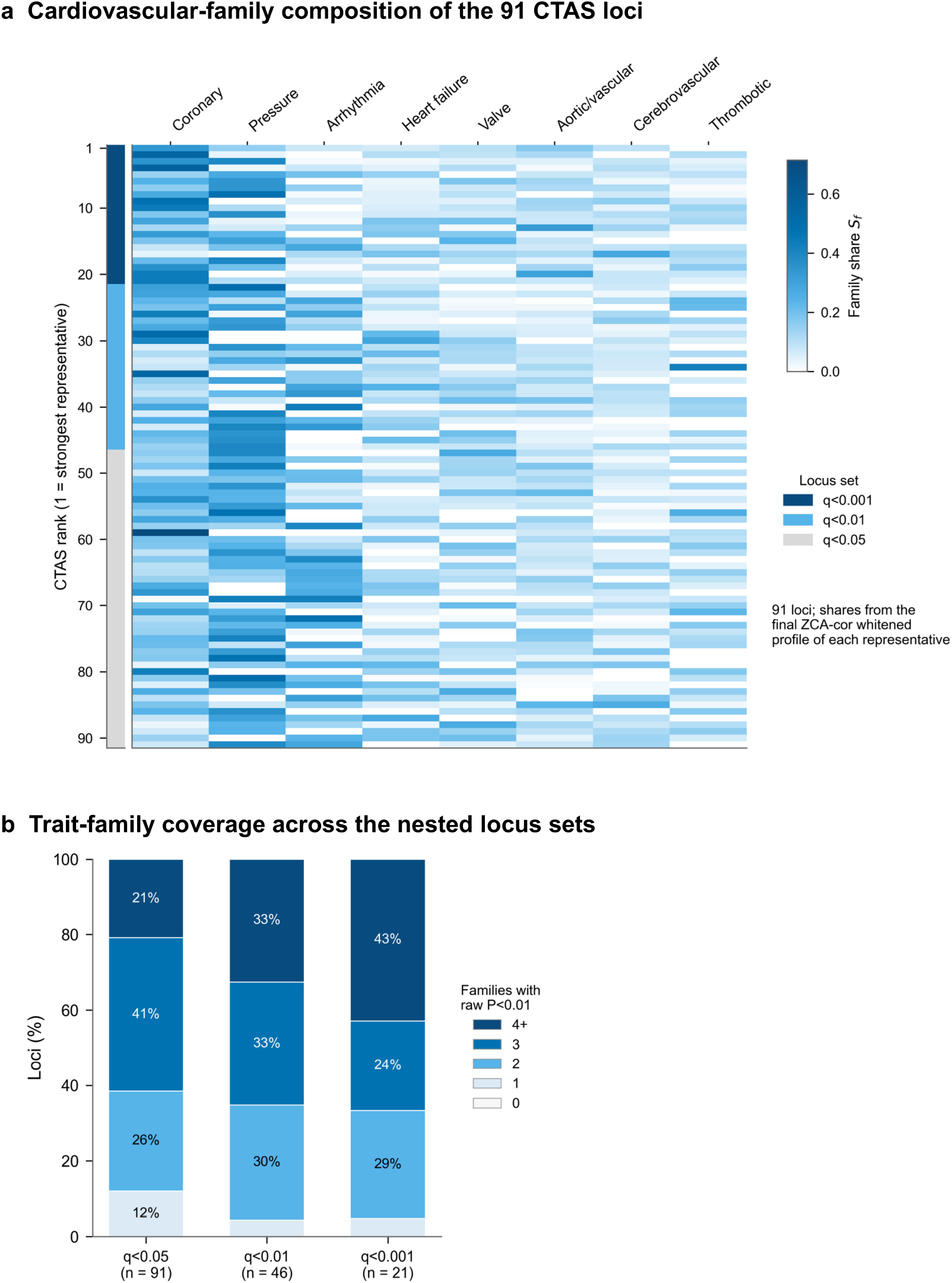
Ranked multi-family architecture of the G1 CTAS loci. (a) Cardiovascular-family composition of all 91 q<0.05 CTAS loci, ordered by the CTAS of each locus’s statistical representative (rank 1 at top; left strip marks the nested q-set). Each row shows the size-adjusted share S_f of positive whitened evidence attributable to each of eight cardiovascular trait families, computed from the representative’s final ZCA-cor whitened profile; coronary is one family among eight. Shares describe the composition of the evidence and are not a selection rule. (b) Descriptive trait-family coverage: the percentage of loci whose representative reaches raw gene-level P<0.01 in 0, 1, 2, 3 or ≥4 of the eight families, for the nested locus sets (q<0.05, n = 91; q<0.01, n = 46; q<0.001, n = 21). Coverage is descriptive only; loci were selected by BH q, not by family coverage. Percentages below 6% are not printed.

### Colocalisation with gene expression

We tested whether association signals at the 91 canonical representative genes were compatible with a shared causal variant for local gene expression and cardiovascular association. Cis-eQTL summary statistics were obtained from seven GTEx datasets [37] in the eQTL Catalogue [38]: heart left ventricle (QTD000256; n=382), atrial appendage (QTD000251; n=372), aorta (QTD000131; n=387), coronary artery (QTD000136; n=213), whole blood (QTD000356; n=670), liver (QTD000266; n=208) and kidney cortex (QTD000261; n=73). For each representative gene, variants within the GRCh38 gene body ±1 Mb were retained when the target Ensembl gene identifier matched, the molecular trait was an SNP and minor-allele frequency exceeded 0.01.

European-ancestry Pan-UK Biobank GWAS effect and quality-control fields (beta_EUR, se_EUR, neglog10_pval_EUR and low_confidence_EUR) were used for all 16 traits in the colocalisation analysis. Estimates were lifted from GRCh37 to GRCh38 and matched to the eQTL data using chromosome, position, reference allele and alternate allele. Direct, allele-swapped, strand-complement and swapped-complement matches were permitted, with effect estimates and allele frequencies flipped where required. Low-confidence GWAS records were excluded, and where duplicate records remained the variant with the smallest GWAS P value was retained. GWAS allele frequency was calculated as the case–control sample-size-weighted European allele frequency. A locus–tissue–trait combination was analysed only when at least 50 variants were matched.

Colocalisation was performed using coloc.abf in coloc v5.2.3 [39, 40]. GWAS data were specified as case–control with the corresponding case fraction, and eQTL data as quantitative. Prior probabilities were *p*_1_ = 10^−4^, *p*_2_ = 10^−4^ and *p*_12_ = 10^−5^. We defined PP4≥0.5 as colocalisation support and PP4≥0.8 as strong support. A locus was classified as PP3-only when at least one evaluable test had PP3≥0.5 but none had PP4≥0.5. Genes whose bodies lay in the extended major histocompatibility complex (GRCh38 chromosome 6: 28.5–33.5 Mb) were not assessed, and genes without a qualifying cis-eQTL were classified as not assessable rather than negative (Fig. 4; Supplementary Fig. 4; Supplementary Table 9).

**Figure 4.**
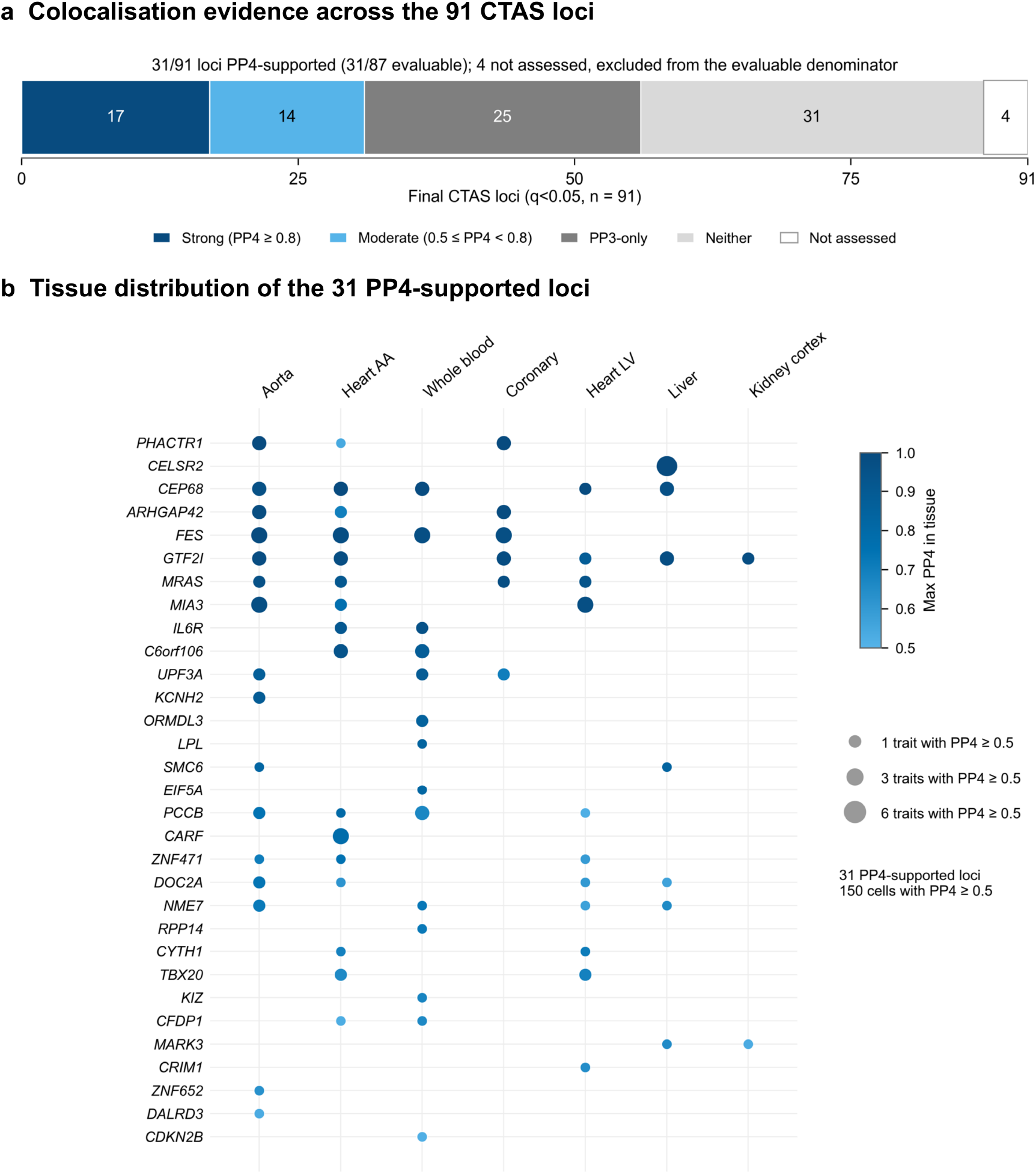
Colocalisation evidence for the G1 CTAS loci. (a) Locus-level colocalisation classes for the statistical representative gene of each final q<0.05 locus, using 7 GTEx cis-eQTL tissues and 16 cardiovascular traits (coloc.abf): 17 strong (maximum PP4 ≥ 0.8), 14 moderate (0.5 ≤ PP4 < 0.8), 25 PP3-only (evidence favouring distinct variants), 31 neither and 4 not assessed. PP4 support (PP4 ≥ 0.5) is seen at 31/91 loci overall and 31/87 evaluable loci; the 4 not-assessed loci (MHC exclusion, or no cis-eQTL for the representative gene) are excluded only from the evaluable denominator and are not negative results. (b) Tissue distribution of the 31 PP4-supported loci (rows, ordered by maximum PP4). Point size is the number of traits with PP4 ≥ 0.5 in that tissue and colour is the maximum PP4 in that tissue; 150 locus × tissue × trait cells reach PP4 ≥ 0.5. Colocalisation supports a shared-variant model for the tested representative-gene eQTL and cardiovascular association; it does not establish the representative gene as causal. AA, atrial appendage; LV, left ventricle.

### Comparison with previous multi-trait studies

Convergent support was assessed against four fixed cardiovascular studies that differed in analytical method and reporting unit. The 38 multi-trait cardiovascular loci reported by Qiao et al. [8] were matched by direct overlap between their GRCh37 intervals and the CTAS locus intervals, without an added flank. The 180 unique genes reported by Liu et al. [9] were matched by HGNC-normalised gene symbol to any q<0.05 CTAS gene within each locus. Coronary heart disease colocalisation regions reported by Foley et al. [18] were matched by the GRCh37 coordinate of their candidate lead variant falling within a CTAS locus. The 625 approximately independent PLEIO index variants reported by Lee et al. [23] were matched to CTAS loci by the same coordinate rule. All four studies were harmonised to the locked CTAS GRCh37/hg19 locus map, their differing native units (region, gene and variant) retained and documented, and reciprocal CTAS-locus recovery was evaluated across the nested 91/46/21 locus sets. Support was summarised for each source separately and for their union. Because phenotype coverage, methods and matching units differed between studies, these comparisons were interpreted as literature convergence, not formal replication (Fig. 5; Supplementary Fig. 5; Supplementary Table 10).

**Figure 5.**
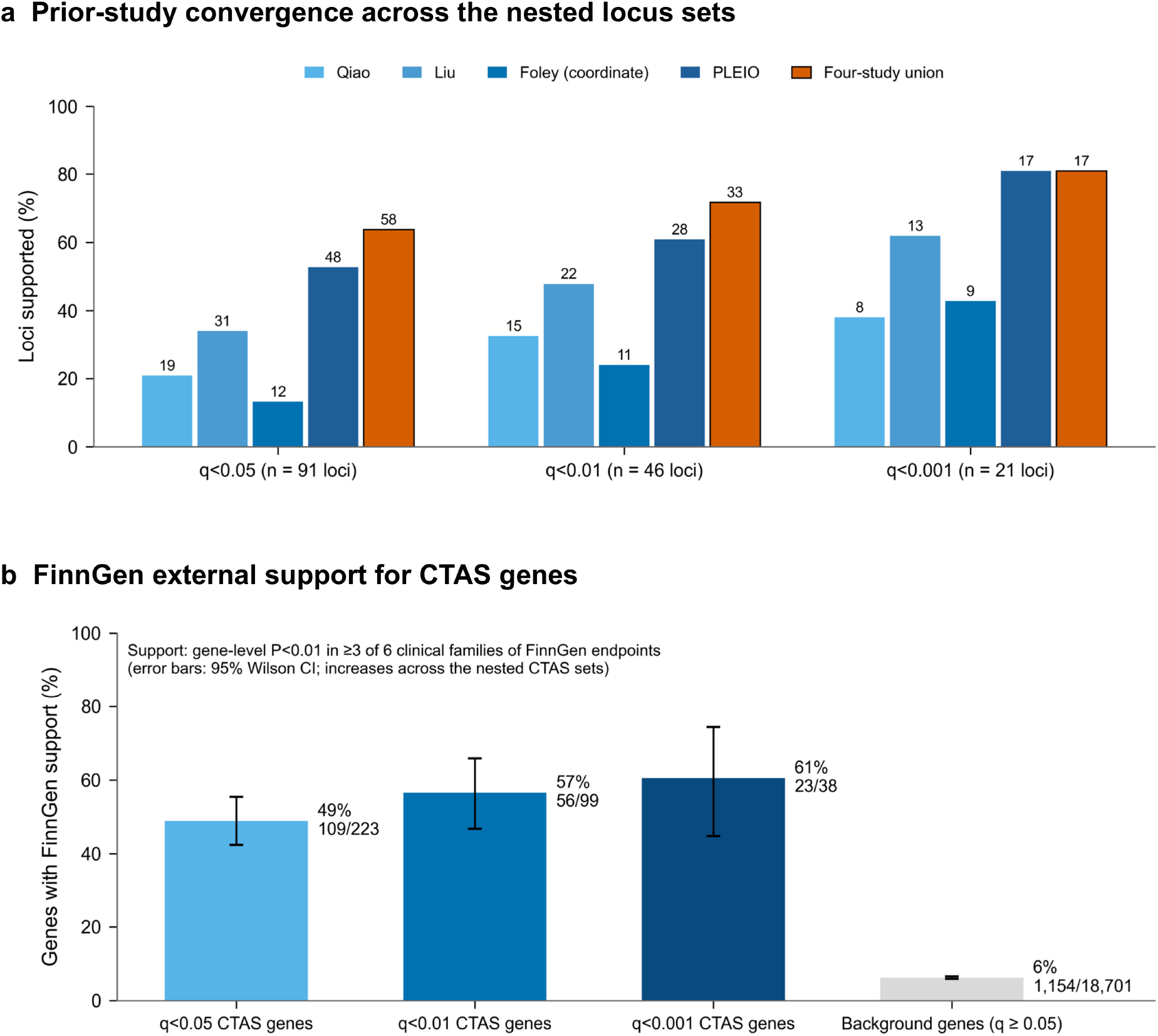
External support for the CTAS loci and genes. (a) Convergent prior-study support across the nested CTAS locus sets: percentage of loci represented in Qiao, Liu, Foley (candidate-variant coordinate containment) and PLEIO, and in their union, each matched on its native unit. Counts above bars: q<0.05 (91 loci) 19 / 31 / 12 / 48 / 58; q<0.01 (46) 15 / 22 / 11 / 28 / 33; q<0.001 (21) 8 / 13 / 9 / 17 / 17 (Qiao / Liu / Foley / PLEIO / union). This is convergence with prior cross-trait resources, not independent confirmation; loci not represented in any comparator are not thereby novel. (b) FinnGen external support for CTAS genes (gene unit): percentage of evaluable genes with gene-level MAGMA P<0.01 in at least 3 of 6 clinical families of FinnGen R12 cardiovascular endpoints (coronary, arrhythmia, myocardial, pressure, cerebrovascular, venous; a family is supported if any member endpoint reaches P<0.01), with 95% Wilson confidence intervals. q<0.05: 109/223 (49%); q<0.01: 56/99 (57%); q<0.001: 23/38 (61%); background genes with q ≥ 0.05: 1,154/18,701 (6%). Support increases across the nested sets, which are not independent groups. Genes absent from the FinnGen outputs are not evaluable and are excluded from denominators. FinnGen is independent-cohort external support for UK Biobank-selected CTAS genes; CTAS was not rerun in FinnGen, so this does not re-estimate the statistic and does not show that the correlation transformation or breadth term is superior.

### FinnGen external-support analysis

This external-support analysis tested whether CTAS-prioritised genes were enriched for independent cardiovascular gene-level evidence in FinnGen. CTAS was not recalculated in FinnGen; instead, the analysis evaluated the transfer of the discovery-derived prioritisation to an external cohort with related cardiovascular endpoints. This provided related-endpoint support rather than exact replication of the 16-trait Pan-UKBB analysis.

Independent cardiovascular support was evaluated using FinnGen release 12 [41] for ten endpoints: atrial fibrillation (I9_AF), angina (I9_ANGINA), coronary atherosclerosis (I9_CORATHER), cardiomyopathy (I9_CARDMYO), heart failure (I9_HEARTFAIL), hypertension (I9_HYPTENS), myocardial infarction (I9_MI_STRICT), stroke (I9_STR), transient ischaemic attack (I9_TIA) and venous thromboembolism (I9_VTE). The primary unit was the gene: every CTAS gene in each nested set was evaluated, and genes absent from the FinnGen gene outputs were classified as not evaluable rather than unsupported. The background comprised genes with q≥0.05 present in the union of the ten FinnGen datasets. A companion analysis evaluated one statistical locus representative per CTAS locus.

FinnGen GWAS summary statistics were obtained on GRCh38 and converted to GRCh37/hg19 using the UCSC hg38ToHg19 chain [42] applied through a validated Python implementation of the chain mapping (the UCSC liftOver binary was not used) before gene annotation. Converted positions were validated against the 1000 Genomes European reference (99.99% agreement among uniquely matched rsIDs), and variants that could not be converted (0.9%) were excluded. Gene-level P values were calculated separately for each endpoint using MAGMA v1.10, the SNP-wise mean model, NCBI build 37.3 gene annotation, a gene-body annotation window with no additional flank and the European 1000 Genomes linkage-disequilibrium reference panel. A uniform sample-size parameter of 500,348 was specified across the ten FinnGen analyses. These internally generated MAGMA outputs formed the gene-level external-support matrix used to evaluate the discovery-derived CTAS genes and loci.

Ten FinnGen endpoints were grouped into six clinical families: coronary (angina, coronary atherosclerosis, myocardial infarction), arrhythmia (atrial fibrillation), myocardial (cardiomyopathy, heart failure), pressure (hypertension), cerebrovascular (stroke, transient ischaemic attack) and venous (venous thromboembolism). The FinnGen venous thromboembolism endpoint (I9_VTE) is defined independently by FinnGen and is distinct from the Pan-UK Biobank ICD-10 I82 trait. Endpoints were grouped into six clinical families so that correlated endpoints (e.g. CAD, MI and angina) were not counted as independent support. A family was supported for a gene if any of its endpoints had gene-level P<0.01. Breadth of FinnGen support was defined as the number of supported families for each CTAS gene and background gene. The primary summary was support in at least three of the six families; support in at least one and at least two families, the mean number of supported families and support in at least two non-coronary families were also reported. The endpoint-based counts (at least one, two and three of ten endpoints) were retained as a sensitivity analysis (Supplementary Table 11). For each nested CTAS set and family-count threshold, enrichment relative to background was evaluated using one-sided Fisher exact tests, with Woolf 95% confidence intervals for odds ratios and Wilson 95% confidence intervals for proportions. Benjamini–Hochberg correction was applied across the nine combinations of three CTAS sets and three family-count thresholds. Because the CTAS sets are nested, they are not independent groups, so disjoint q-value bands were also compared descriptively. The same summaries were calculated for the locus-representative companion analysis (Fig. 5; Supplementary Fig. 5; Supplementary Table 11). In the evidence integration of Table 3, FinnGen support for a locus representative was defined as gene-level P<0.01 in at least one endpoint.

**Table 3.** Integrated evidence across the stringent CTAS locus set. The complete set of 21 canonical loci surviving genome-wide Benjamini–Hochberg q<0.001, one row per locus in descending CTAS of the statistical representative; inclusion is fixed by the prespecified q<0.001 threshold, and the downstream evidence streams never determine which loci appear. Each row gives the statistical representative gene (the highest-CTAS significant member; internal locus identifiers are listed in Supplementary Table 3); CTAS with its BH q-value from empirical P values over 10^9^ null draws, and the magnitude M and effective breadth PR that compose it (CTAS = M × log2(1 + PR)); the representative’s raw UK Biobank signal profile, listing the named traits at gene-level P<0.01 in Table 2 panel order; a separate descriptive cross-family summary computed with the final ZCA-cor whitening (Family N_eff_ and the leading family, with two co-leading families shown when the second share is at least 90% of the first); the strongest colocalisation reaching PP4≥0.5 with its tissue and trait, with “No PP4 support” and “Not assessed” (ATP6V1G2, MHC exclusion) kept distinct; representative-gene FinnGen support, given as the number of six clinical families (and of ten FinnGen endpoints) with gene-level P<0.01 and naming the supported families; representation in the Qiao, Liu, Foley and PLEIO comparator studies, matched on each study’s native unit, with 17 of the 21 core loci supported by at least one study; and a short locus-specific biological-context label. The locus-specific literature for CFHR3, C6orf106, LPL, HTRA1 and LDLR is cited with that for the other core loci. Statistical representatives are reproducible locus labels, not causal-gene assignments, and the absence of support in any single downstream stream (colocalisation, FinnGen or comparator) is complementary information rather than evidence against the locus. Definitions, thresholds and abbreviations are given in the table footnote.

| Representative | CTAS (BH q) | UKBB signal profile | Cross-family profile | Best colocalisation (PP4≥0.5) | FinnGen families / 6 (endpoints / 10) | Prior multi-trait | Literature context |
|---|---|---|---|---|---|---|---|
| **LPA** | **31.16**<br>(8.5×10<sup>−8</sup>) | M 11.14 · PR 5.95<br>8 traits P<0.01: CAD, MI, Ang, HTN, I73, TIA, AoAn, AoV | Family Neff 5.04<br>Leading: coronary | No PP4 support | **1/6 (3/10)**<br>Coronary | Qiao·Liu·Foley·PLEIO | **Anchor** · Lp(a) biology; human genetics support causal coronary risk [43] |
| **CDKN2B** | **26.16**<br>(6.4×10<sup>−6</sup>) | M 11.41 · PR 3.90<br>6 traits P<0.01: CAD, MI, Ang, HF, I73, AoV | Family Neff 2.91<br>Leading: coronary | PP4 0.52<br>whole blood · I80 | **4/6 (7/10)**<br>Coronary, Arrhythmia, Pressure, Cerebrovascular | Qiao·Liu·PLEIO | **Anchor** · 9p21 cell-cycle and efferocytosis biology in atherosclerosis [44] |
| **FES** | **24.27**<br>(7.6×10<sup>−6</sup>) | M 9.72 · PR 4.65<br>4 traits P<0.01: CAD, MI, Ang, HTN | Family Neff 3.39<br>Leading: pressure | PP4 0.99<br>aorta · Ang | **4/6 (6/10)**<br>Coronary, Arrhythmia, Pressure, Cerebrovascular | Liu·Foley·PLEIO | **Functional** · 15q26 CAD-locus kinase; protective in atherosclerosis models [45] |
| **APOE** | **24.26**<br>(7.6×10<sup>−6</sup>) | M 10.98 · PR 3.63<br>4 traits P<0.01: CAD, MI, Ang, AoV | Family Neff 2.80<br>Leading: coronary | No PP4 support | **5/6 (7/10)**<br>Coronary, Myocardial, Pressure, Cerebrovascular, Venous | Qiao·Liu·Foley·PLEIO | **Anchor** · Canonical lipid-transport locus; APOE genotypes associate with lipid levels and coronary risk [46] |
| **MARK3** | **22.46**<br>(2.4×10<sup>−5</sup>) | M 7.57 · PR 6.81<br>7 traits P<0.01: CAD, Ang, AF, CMP, HTN, Ath, I80 | Family Neff 5.74<br>Leading: pressure | PP4 0.65<br>liver · CAD | **3/6 (4/10)**<br>Coronary, Pressure, Cerebrovascular | None of 4 | **Unresolved** · Kinase with metabolic and hepatic biology; cardiovascular mechanism unestablished [47] |
| **CFDP1** | **21.38**<br>(1.3×10<sup>−4</sup>) | M 8.48 · PR 4.74<br>7 traits P<0.01: CAD, MI, Ang, HTN, I73, AoAn, AoV | Family Neff 4.31<br>Leading: pressure | PP4 0.66<br>whole blood · MI | **5/6 (7/10)**<br>Coronary, Arrhythmia, Myocardial, Pressure, Venous | Liu·PLEIO | **Functional** · BCAR1–CFDP1–TMEM170A carotid-IMT/CAD locus; CFDP1 supports cardiac development and function, with BCAR1 the adjacent effector candidate [48,49] |
| **ATP6V1G2** | **21.16**<br>(1.9×10<sup>−4</sup>) | M 8.08 · PR 5.15<br>5 traits P<0.01: CAD, Ang, AF, HTN, I73 | Family Neff 4.99<br>Leading: pressure | Not assessed | **5/6 (7/10)**<br>Coronary, Arrhythmia, Myocardial, Pressure, Venous | Liu·PLEIO | **Unresolved** · V-ATPase subunit within the MHC/6p21 multi-trait cardiovascular–immune region; no single-gene CV effector established (extended LD) |
| **ZNF652** | **20.79**<br>(2.6×10<sup>−4</sup>) | M 9.63 · PR 3.47<br>5 traits P<0.01: CAD, MI, Ang, HTN, AoV | Family Neff 3.11<br>Leading: pressure | PP4 0.63<br>aorta · HTN | **3/6 (5/10)**<br>Coronary, Arrhythmia, Pressure | Qiao·Liu·Foley·PLEIO | **Human-genetic** · ZNF652-region association with blood pressure and hypertension [50] |
| **CARF** | **20.66**<br>(3.0×10<sup>−4</sup>) | M 9.18 · PR 3.76<br>5 traits P<0.01: CAD, MI, Ang, IS, AoAn | Family Neff 3.43<br>Leading: coronary | PP4 0.78<br>atrial appendage · MI | **2/6 (4/10)**<br>Coronary, Cerebrovascular | Liu·PLEIO | **Unresolved** · Statistical representative at the 2q33 WDR12-region coronary/MI locus; CARF (calcium-responsive regulator) has limited direct CV validation [51] |
| **CELSR2** | **20.61**<br>(3.0×10<sup>−4</sup>) | M 9.47 · PR 3.52<br>4 traits P<0.01: CAD, MI, Ang, HTN | Family Neff 3.92<br>Leading: coronary | PP4 1.00<br>liver · CAD | **2/6 (4/10)**<br>Coronary, Arrhythmia | Qiao·Liu·Foley·PLEIO | **Anchor** · 1p13 CELSR2–PSRC1–SORT1 liver-expression CAD/LDL locus; CELSR2 regulates hepatocyte lipid accumulation, SORT1/PSRC1 the adjacent effectors [52,53] |
| **ACAD10** | **20.50**<br>(3.0×10<sup>−4</sup>) | M 8.97 · PR 3.87<br>5 traits P<0.01: CAD, MI, HTN, I73, I80 | Family Neff 4.83<br>Leading: pressure | No PP4 support | **4/6 (6/10)**<br>Coronary, Pressure, Cerebrovascular, Venous | Qiao·Liu·Foley·PLEIO | **Human-genetic** · 12q24 (SH2B3/ALDH2) blood-pressure-associated region; ACAD10 variant associated with sex-specific hypertension susceptibility [54] |
| **HNF1A** | **20.39**<br>(3.4×10<sup>−4</sup>) | M 7.92 · PR 4.95<br>6 traits P<0.01: CAD, MI, Ang, HF, I80, MV | Family Neff 5.37<br>Leading: coronary | No PP4 support | **3/6 (4/10)**<br>Coronary, Arrhythmia, Pressure | Liu·Foley·PLEIO | **Anchor** · Hepatic regulator; coding variants associated with multiple cardiovascular-risk phenotypes [55] |
| **IREB2** | **20.31**<br>(3.6×10<sup>−4</sup>) | M 7.21 · PR 6.04<br>5 traits P<0.01: MI, HF, Ath, I73, AoAn | Family Neff 5.28<br>Leading: aortic/vascular | No PP4 support | **2/6 (3/10)**<br>Coronary, Pressure | PLEIO | **Unresolved** · Iron-response regulator in the 15q25 region that also carries the established ADAMTS7 coronary signal; the CTAS signal is not equated with ADAMTS7 [56] |
| **TBX20** | **20.02**<br>(4.4×10<sup>−4</sup>) | M 7.10 · PR 6.07<br>6 traits P<0.01: CAD, MI, Ang, HF, HTN, AoV | Family Neff 4.42<br>Leading: coronary | PP4 0.69<br>left ventricle · CAD | **1/6 (1/10)**<br>Coronary | None of 4 | **Functional** · Cardiac transcription factor; human variants associated with developmental heart disease and cardiomyopathy [57,58] |
| **DALRD3** | **19.92**<br>(4.9×10<sup>−4</sup>) | M 6.76 · PR 6.71<br>6 traits P<0.01: CAD, MI, Ang, HTN, AoV, MV | Family Neff 4.42<br>Leading: pressure | PP4 0.53<br>aorta · MV | **3/6 (3/10)**<br>Coronary, Arrhythmia, Pressure | Foley·PLEIO | **Unresolved** · tRNA-modification gene labelling the 3p21.31 coronary-artery-disease region; published signals map near RHOA, without establishing a causal gene [59,60] |
| **ORMDL3** | **19.83**<br>(5.1×10<sup>−4</sup>) | M 6.70 · PR 6.79<br>4 traits P<0.01: AF, HF, HTN, I82 | Family Neff 6.10<br>Leading: arrhythmia | PP4 0.85<br>whole blood · AF | **2/6 (2/10)**<br>Arrhythmia, Myocardial | Liu·PLEIO | **Emerging** · Sphingolipid regulator with endothelial and atherosclerosis evidence [61] |
| **CFHR3** | **19.49**<br>(7.8×10<sup>−4</sup>) | M 6.38 · PR 7.31<br>2 traits P<0.01: IS, TIA | Family Neff 5.67<br>Leading: cerebrovascular | No PP4 support | **0/6 (0/10)** | None of 4 | **Emerging** · Complement factor-H-related gene at 1q31.3 with alternative-complement-pathway biology; no established cardiovascular effector assigned [62] |
| **C6orf106** | **19.40**<br>(8.5×10<sup>−4</sup>) | M 8.16 · PR 4.20<br>4 traits P<0.01: CAD, Ang, HTN, I82 | Family Neff 4.29<br>Leading: pressure | PP4 0.94<br>atrial appendage · HTN | **5/6 (8/10)**<br>Coronary, Arrhythmia, Myocardial, Pressure, Cerebrovascular | Qiao·PLEIO | **Human-genetic** · C6orf106 (ILRUN) labels the 6p21.31 region associated with coronary artery disease; regional effector not established [63] |
| **HTRA1** | **19.38**<br>(8.5×10<sup>−4</sup>) | M 7.50 · PR 4.99<br>4 traits P<0.01: CAD, MI, Ang, HTN | Family Neff 4.41<br>Leading: coronary | No PP4 support | **2/6 (3/10)**<br>Coronary, Pressure | None of 4 | **Human-genetic** · Vessel-wall serine protease; loss of function causes CARASIL, and variants associate with cerebral small-vessel disease and stroke [64,65] |
| **LDLR** | **19.33**<br>(9.0×10<sup>−4</sup>) | M 7.63 · PR 4.78<br>4 traits P<0.01: CAD, MI, Ang, I73 | Family Neff 3.31<br>Leading: coronary | No PP4 support | **4/6 (7/10)**<br>Coronary, Arrhythmia, Pressure, Cerebrovascular | Qiao·Liu·Foley·PLEIO | **Anchor** · Hepatic LDL receptor; familial hypercholesterolaemia gene and canonical coronary-atherosclerosis locus [66] |
| **LPL** | **19.25**<br>(9.8×10<sup>−4</sup>) | M 8.46 · PR 3.84<br>4 traits P<0.01: CAD, MI, Ang, HTN | Family Neff 4.15<br>Leading: coronary | PP4 0.84<br>whole blood · Ang | **4/6 (6/10)**<br>Coronary, Arrhythmia, Myocardial, Pressure | PLEIO | **Anchor** · Lipoprotein lipase; established determinant of plasma triglycerides and coronary artery disease [67] |

The complete set of 21 canonical loci surviving genome-wide Benjamini–Hochberg q<0.001, ordered by descending CTAS of the statistical representative; downstream evidence did not determine inclusion. Statistical representatives label canonical loci and are not causal-gene assignments. CTAS, Cross-Trait Aggregation Score; BH q, Benjamini–Hochberg q-value (empirical P from 10<sup>9</sup> null draws). CTAS is computed jointly from all 16 gene-level trait signals after ZCA-cor whitening with the transformed trait correlation matrix R = 2 sin(πρS/6), as CTAS = M × log2(1 + PR). UKBB signal profile: M is the magnitude of the positive whitened evidence and PR its participation ratio across the positive whitened axes; because those axes are mixtures of the original traits, PR is not a count of named traits. The named UKBB traits shown are those with raw gene-level P<0.01 (Table 2 panel order) and are descriptive, not additive components of CTAS. Cross-family profile: a separate descriptive summary of the same whitened evidence — Family Neff, the effective number of contributing cardiovascular trait families across eight named families, and the leading family (two co-leading families are shown when the second-largest size-adjusted share is at least 90% of the largest). Best colocalisation: maximum PP4 shown only when PP4≥0.5, with GTEx tissue and trait; “No PP4 support” denotes PP3-only or neither class and is kept distinct from “Not assessed” (MHC exclusion or no cis-eQTL). PP4 supports a shared-variant model, not a causal gene. FinnGen: number of six clinical families (coronary: Ang, CAD, MI; arrhythmia: AF; myocardial: CMP, HF; pressure: HTN; cerebrovascular: Stroke, TIA; venous: VTE, the FinnGen I9_VTE endpoint) with gene-level P<0.01 in at least one member endpoint, supported families named, followed in brackets by the number of the ten FinnGen R12 endpoints with P<0.01; endpoints were grouped so that correlated endpoints are not counted as independent support. All 21 representatives were evaluable and P values are from the corrected GRCh37 analysis (4 November 2026). CTAS was not rerun in FinnGen, so this is independent-cohort external support. Prior multi-trait: representation in four prior cross-trait cardiovascular resources — Qiao, Liu, Foley and PLEIO — each matched on its native unit against the canonical GRCh37/hg19 loci (Qiao region and Foley candidate-variant coordinate overlap; Liu pleiotropic-gene membership; PLEIO index-variant coordinate overlap); “None of 4” = comparator-unmatched, not evidence of novelty or absent biology; this is convergence, not independent confirmation. Literature context is orienting: Anchor, established cardiovascular locus; Functional, experimental support; Human-genetic, human genetic association support; Emerging, emerging evidence; Unresolved, effector biology not yet resolved. Bracketed numbers are manuscript references 43–67. Trait abbreviations: AF, atrial fibrillation; Ang, angina; AoAn, aortic aneurysm; AoV, aortic valve disease; Ath, atherosclerosis; CAD, coronary artery disease; CMP, cardiomyopathy; HF, heart failure; HTN, hypertension; I73, other peripheral vascular disease; I80, phlebitis/thrombophlebitis (complete ICD-10 I80 category); I82, venous embolism/thrombosis (complete ICD-10 I82 category); IS, ischaemic stroke; MI, myocardial infarction; MV, mitral valve disease; TIA, transient ischaemic attack. In the FinnGen column, VTE denotes the separately defined FinnGen venous thromboembolism endpoint (I9_VTE), which is distinct from I82.

### Software and reproducibility

Data preparation and analyses were performed using R 4.5.1, Python 3.13.2 and shell scripts. Core analyses used MAGMA v1.10 for gene-level association and coloc v5.2.3 for colocalisation. Genome-build conversion used UCSC chain files, with the software implementation differing by analysis: for colocalisation, GWAS coordinates were converted from GRCh37 to GRCh38 with the UCSC liftOver command-line program and the hg19ToHg38 chain; for FinnGen, coordinates were converted from GRCh38 to GRCh37 with the hg38ToHg19 chain through a custom Python implementation, and the liftOver program was not used. No other coordinate conversion was applied. Random seeds were fixed in all simulation and resampling analyses as specified above. Primary inputs, analysis scripts, intermediate validation files and final tabular outputs were frozen with the completed analysis package so that reported gene sets, locus assignments and downstream results could be regenerated without manual selection. Data and code availability are described in the Declarations.

### Use of AI tools

Generative AI tools (ChatGPT, OpenAI; Claude, Anthropic) were used during manuscript and software development for language editing, code assistance and quality-control support. All analyses, code, figures, factual claims and interpretations were reviewed and verified by the authors, who take full responsibility for the work. Generative AI was not used to generate or alter primary data.

## Results

### CTAS provides calibrated gene-level evidence across correlated cardiovascular traits

To distinguish broadly shared genetic association from recurrence expected between correlated cardiovascular traits, we applied CTAS to gene-level association profiles across the 16-trait panel. Gene-level signals were correlated across traits: off-diagonal entries of the transformed rank-correlation matrix ranged from −0.019 to 0.462, and its minimum eigenvalue was 0.50 (Fig. 1a). We therefore decorrelated each gene’s association profile by ZCA-cor whitening before calculating CTAS, which combines the magnitude and breadth of positive evidence after dependence correction. CTAS breadth reflects evidence distributed across decorrelated axes, not a simple count of associated traits.

We calibrated each gene’s CTAS against an empirical null of 10^9^ profiles simulated from the fitted multivariate normal model and adjusted the resulting empirical P values across 19,080 genes using the Benjamini–Hochberg procedure. This identified 223 genes mapping to 91 loci at q<0.05, with nested sets of 99 genes at 46 loci at q<0.01 and 38 genes at 21 loci at q<0.001 (Fig. 1b; Supplementary Tables 1–3). Gene and locus membership was identical across null seeds 42, 1 and 2. These loci do not bypass genome-wide multiple-testing control: significance is assigned to the prespecified cross-trait gene-level hypothesis across all 19,080 tested genes, and does not require an individual component trait to cross a single-trait significance threshold.

LPA had the highest CTAS (31.16; empirical P=4.47×10^−12^, an upper-tail estimate; q=8.52×10^−8^), followed by CDKN2B, while FES and APOE were also among the highest-ranking locus representatives (Fig. 1b). The highest-ranking signals were distributed across the autosomes, not concentrated in a single genomic region. Locus representatives denote the highest-CTAS gene within each locus and are not, by themselves, causal-gene assignments. The resulting 91-locus set was carried forward for robustness testing, analysis of cross-trait signal, colocalisation and assessment of external support. As an illustration, DALRD3 entered the most stringent q<0.001 core despite no individual trait reaching gene-wide significance (lowest single-trait gene-level P=4.5×10^−5^); its evidence instead comprised coordinated subthreshold association across six traits, with additional support from colocalisation, a FinnGen association (P<0.01 in at least one endpoint) and prior multi-trait studies (Table 3).

### CTAS is calibrated and stable across analytical perturbations

We next evaluated whether CTAS was calibrated under its fitted null and behaved as intended. Using 200,000 independently simulated null genes evaluated against a 1,000,000-draw reference null, observed type-I error closely followed nominal probability, including 0.050 at α=0.05, 0.0099 at α=0.01 and 0.00088 at α=0.001 (Fig. 2a; Supplementary Table 4). Under the primary 10^9^-draw null, 220 of the 223 q<0.05 genes had directly counted empirical P values; two exceeded every null draw and took the floor value, and one (LPA) received an upper-tail estimate (Supplementary Figs. 1 and 2; Supplementary Table 2). Because the empirical P values at the q-value boundaries were far above the resolution limit, tail handling did not change the q-value set assigned to any gene.

When a fixed total simulated signal was distributed across increasing numbers of decorrelated axes, CTAS increased (Fig. 2b; Supplementary Fig. 2; Supplementary Table 4), showing that the breadth term captures signal spread. In a sensitivity analysis, replacing the transformed rank-correlation matrix with the raw Spearman matrix left genome-wide gene ordering essentially unchanged (Spearman ρ=0.99996), and every primary gene and locus set was contained in the corresponding sensitivity set (gene Jaccard 0.93, 0.93 and 0.84 at q<0.05, q<0.01 and q<0.001; Fig. 2c; Supplementary Table 5). The prespecified primary CTAS supplied the FDR-controlled gene sets used in every downstream analysis.

The locus set was also stable across analytical perturbations. These sensitivity analyses used their recorded raw Spearman working correlation matrices and 10^6^-draw nulls, and their results were evaluated against the primary gene and locus sets (Methods). Replacing the common MAGMA sample-size parameter with trait-specific effective sample sizes left the gene ranking essentially unchanged (Spearman ρ=0.999998) and changed the status of none of the 223 genes or 91 loci at any q-value threshold (Supplementary Table 6). Across 16 leave-one-trait-out analyses, 47 of 91 loci were retained after every omission, with a mean of 80.1 loci retained per analysis (range, 67–89; Supplementary Fig. 2; Supplementary Table 7). In a more stringent 10-trait analysis that removed six overlapping coronary, peripheral vascular and cerebrovascular manifestations and retained coronary artery disease as the sole representative of the atherosclerotic and obstructive cluster, 65 of 91 q<0.05 loci and 27 of 46 q<0.01 loci were regionally retained. Of the 21 core loci, 16 were represented at reduced-panel q<0.05 and 12 at q<0.01, although none reached q<0.001 (Supplementary Table 8). This reduced panel is a deliberately stringent sensitivity analysis. By collapsing the correlated obstructive and atherosclerotic phenotypes to coronary artery disease alone and removing the cerebrovascular manifestations, it tests whether the result is an artefact of jointly including several closely related obstructive, atherosclerotic and cerebrovascular manifestations. Retention of a substantial locus core under this test indicates that the reported loci do not simply reflect the same coronary signal measured repeatedly. These analyses show that the CTAS loci are internally stable across analytical perturbations.

### CTAS loci form a ranked, multi-family cardiovascular architecture

The 91 prioritised loci showed distinct combinations of association evidence across clinically coherent cardiovascular families (Fig. 3a; Supplementary Fig. 3a; Supplementary Table 3). At q<0.05, 88% of loci had representative-gene associations spanning at least two families. The proportion spanning four or more families increased across the nested CTAS tiers, from 21% at q<0.05 to 33% at q<0.01 and 43% at q<0.001 (Fig. 3b). The effective number of contributing families (Family N_eff_) had a median of 4.57 across the 91 loci (interquartile range, 3.78–5.27; range, 1.88–7.28). Family coverage was calculated descriptively from the canonical locus representative and was not used to select loci. These clinical-family profiles complement CTAS breadth by resolving the composition of evidence across named disease groups.

We next characterised the organisation of these family profiles in multivariate space and assessed whether they supported stable discrete locus groupings. Ordination of the size-adjusted family contributions separated broad compositional tendencies across the first two components, which explained 28.2% and 25.9% of the variance, respectively (Supplementary Fig. 3b; Supplementary Table 3). However, cluster separation was weak across the tested solutions, with a best mean partitioning-around-medoids silhouette of 0.185 (k=3; bootstrap stability 0.66). The CTAS loci therefore formed an overlapping multi-family continuum, not discrete classes, with the highest-confidence loci increasingly supported across multiple families.

### CTAS loci show tissue-specific colocalisation with gene expression

To investigate whether prioritised CTAS signals shared genetic variants with local gene regulation, we tested the canonical representative gene at each of the 91 loci across seven GTEx cis-eQTL datasets and the 16 cardiovascular traits. Across the complete set of 91 loci, 31 showed colocalisation support (maximum PP4≥0.5): 17 had strong support (PP4≥0.8) and 14 had moderate support (0.5≤PP4<0.8); 87 loci were evaluable and four were not assessed (Fig. 4a; Supplementary Table 9). The proportion of supported loci increased across the nested CTAS sets, from 31 of 91 at q<0.05 to 22 of 46 at q<0.01 and 12 of 21 at q<0.001 (31 of 87, 22 of 44 and 12 of 20 evaluable loci).

Colocalisation was tissue-specific and distributed across all seven datasets (Fig. 4b; Supplementary Fig. 4a). Aorta contributed the largest number of supported locus–tissue–trait combinations, followed by atrial appendage and whole blood, although support was not restricted to vascular tissues. Strong examples included PHACTR1 in aorta and coronary artery, CELSR2 in liver, CEP68 in atrial appendage and FES in aorta. Of the 10,192 locus–tissue–trait combinations, 9,056 were assessed; 150 reached PP4≥0.5 and 83 reached PP4≥0.8, showing that supported loci often connected to more than one cardiovascular trait or tissue.

Among the remaining loci, 25 showed PP3-only evidence, consistent with distinct association and expression variants within the same region, while 31 were evaluable but reached neither posterior-probability threshold (Supplementary Fig. 4b–d; Supplementary Table 9). Four loci were not assessed: ATP6V1G2 was excluded because its locus lies within the MHC region, while HOXD13, HDGFL1 and KCNK4 lacked a qualifying cis-eQTL and were not treated as negative results. The ATP6V1G2 label denotes the statistical representative of a gene-dense extended MHC/6p21 interval whose CTAS-significant members include BAG6 and numerous immune-region genes; the dense linkage disequilibrium across this interval precludes clean attribution to a single causal effector, so the appropriate interpretation is a recurrent immune–vascular region, not an ATP6V1G2-specific mechanism. Colocalisation provided tissue-resolved regulatory support for a substantial subset of the CTAS loci while distinguishing these loci from regions with evidence for separate association and expression signals.

### CTAS loci and genes carry convergent and external cardiovascular support

To assess whether CTAS prioritisation recovered loci supported beyond the discovery analysis, we compared the 91 loci with four previous multi-trait cardiovascular studies (Fig. 5a; Supplementary Fig. 5a; Supplementary Table 10). At q<0.05, Qiao, Liu, Foley and PLEIO supported 19, 31, 12 and 48 of the 91 CTAS loci, respectively, with 58 loci (63.7%) represented in at least one study. Union support was 33 of 46 loci (71.7%) at q<0.01 and 17 of 21 loci (81.0%) at q<0.001, and PLEIO alone recovered all 17 of these core loci. Exact one-to-one agreement with these resources is not expected, because they differ in phenotype scope, statistical method, inferential aim, matching unit and significance thresholds; the greater representation among more stringent CTAS loci reflects convergence on shared cardiovascular signal, not replication.

We next evaluated the CTAS genes in FinnGen R12 across ten cardiovascular endpoints grouped into six clinical families. All 223 q<0.05 genes, including all 99 q<0.01 and all 38 q<0.001 genes, were evaluable. The proportion of genes with gene-level P<0.01 in at least three of the six families increased across the nested sets, from 49% at q<0.05 (109 of 223) to 57% at q<0.01 (56 of 99) and 61% at q<0.001 (23 of 38), compared with 6% of evaluable background genes with q≥0.05 (1,154 of 18,701; Fig. 5b; Supplementary Fig. 5b,c; Supplementary Table 11). Each nested set was enriched relative to background (one-sided Fisher exact test; odds ratios 14.5 [95% CI 11.1–19.0], 19.8 [13.2–29.6] and 23.3 [12.1–44.8]; Benjamini–Hochberg-adjusted P≤1.3×10^−18^). Enrichment was also present for support in at least one or at least two families. Disjoint q-value bands were compared descriptively for support in at least three families (Supplementary Table 11): 43% (53 of 124) at 0.01≤q<0.05, 54% (33 of 61) at 0.001≤q<0.01 and 61% (23 of 38) at q<0.001. In a companion analysis of one statistical representative per locus, all 91 representatives were evaluable. Specifically, 42 of 91 (46.2%), 25 of 46 (54.3%) and 13 of 21 (61.9%) had support in at least three families at q<0.05, q<0.01 and q<0.001, respectively. In a sensitivity analysis counting individual endpoints, support in at least three of ten endpoints was 70%, 81% and 89% of CTAS genes at q<0.05, q<0.01 and q<0.001, against 10% of background genes (Supplementary Table 11).

Integrating these evidence streams across the complete 21-locus q<0.001 core revealed complementary rather than redundant support (Table 3). In this core, 12 of the 21 loci (12 of 20 evaluable) had colocalisation support at PP4≥0.5, 20 of the 21 representatives had a FinnGen association (P<0.01 in at least one endpoint), and 17 of the 21 were represented in at least one of four comparator studies. Ten loci (CDKN2B, FES, CFDP1, ZNF652, CARF, CELSR2, DALRD3, ORMDL3, C6orf106 and LPL) had evidence in all three streams. Where available, the literature context shown in Table 3 was drawn from locus-specific genetic and functional studies [43–67].

## Discussion

CTAS identified 223 genes at 91 loci across 16 correlated cardiovascular traits, including a stringent core of 38 genes at 21 loci. Some of these signals may be missed by significance-thresholded single-trait discovery: DALRD3 entered the core without any single trait reaching gene-level significance, and MMP16 had the most distributed profile of any locus. The prioritised loci spanned multiple clinical families, about one third colocalised with local gene expression, the most stringent were increasingly represented in previous multi-trait studies, and CTAS genes were enriched about eight-to ten-fold for support across FinnGen clinical families. A complete correlated panel can therefore be analysed once, with dependence discounted inside the statistic, to give calibrated and externally supported gene-level priorities.

CTAS draws on established methods for rank-based dependence estimation, symmetric (ZCA-cor) whitening, effective-dimensionality measurement, Monte Carlo calibration and false-discovery control. Its contribution is to combine them into a single gene-level statistic that evaluates the complete correlated panel once, with dependence correction inside the score and both the magnitude and spread of residual evidence retained. This targets a question that existing multi-trait methods do not directly address: which genes carry the strongest positive association profiles once the recurrence expected from panel-wide dependence has been discounted? Other methods borrow information to increase trait-specific power, search for supported phenotype subsets, test pairwise pleiotropy, model latent genetic structure or evaluate shared causal variants (Table 1) [13]. Omnibus multi-trait gene and SNP-set tests [20–22] summarise multivariate association as a single P value; CTAS instead makes the magnitude and breadth of dependence-adjusted positive evidence the object of inference. Excess cross-trait evidence of this kind is consistent with shared cardiovascular biology but does not by itself establish causal pleiotropy or a shared mechanism.

The simulation and sensitivity analyses support the statistical validity and robustness of CTAS. Type-I error followed its nominal level under an independently simulated null, while the fixed-total-signal experiment showed that CTAS increases as a fixed total signal is spread across more decorrelated axes. Replacing the transformed rank-correlation matrix with the raw Spearman matrix barely changed the genome-wide ordering, and primary membership was identical across null seeds. Stability was reinforced by the effective-sample-size analysis, 16 leave-one-trait-out analyses and the deliberately stringent reduced-panel analysis, each evaluated against the primary gene and locus sets, which retained substantial portions of the locus set despite changing the input panel.

Biologically, the shared cardiovascular signal was broad and overlapping, not modular. Most loci had evidence spanning more than one clinical family, and coverage of four or more families became more common at greater stringency, so the strongest cross-trait signals increasingly crossed conventional clinical boundaries. The loci carried different mixtures of coronary, vascular, cardiac, valvular, cerebrovascular, pressure and thrombotic evidence, consistent with shared processes distributed across anatomical and clinical compartments [4, 8].

Colocalisation provided a tissue-resolved layer of regulatory support for the prioritised loci. About one third of the 91 loci (31 of 91; 31 of 87 evaluable) had evidence compatible with a shared causal variant between cardiovascular association and local gene expression, and supported locus–tissue–trait combinations were distributed across all seven expression datasets. The prominence of aorta and atrial appendage was biologically plausible, while support in blood, liver and other tissues showed that the regulatory landscape was not confined to a single cardiovascular compartment [37, 59]. These results connect a substantial subset of prioritised loci to plausible regulatory contexts and nominate specific locus–tissue–trait combinations for mechanistic follow-up. PP4 supports a shared-variant model, not the identity of the causal gene or variant [39], so colocalisation refines prioritisation without replacing functional validation.

Two external evidence streams indicate that CTAS concentrates cardiovascular signal with relevance beyond the discovery data. Representation in four previous multi-trait studies increased across the nested sets, reaching 17 of the 21 core loci despite differences in phenotype composition, statistical unit and design [8, 9, 18, 23]. In FinnGen, which was designed as an external gene-level support analysis and not as a reconstruction of CTAS, support across at least three of six clinical families rose across the nested sets, from about eight times the background rate at q<0.05 to about ten times at q<0.001 [41]. Part of any gradient across thresholds is expected, because greater stringency selects stronger signals; the more informative findings are the enrichment over background in an independent cohort and the convergence with heterogeneous published analyses.

Integrating the evidence streams across the stringent q<0.001 core clarifies how these analyses complement one another (Table 3). Established cardiovascular anchors such as LPA, APOE and CELSR2 sit alongside loci with complementary or differing patterns of support: MARK3 and TBX20 are unmatched by the four comparator studies yet show tissue-specific colocalisation (in liver and left ventricle, respectively). IREB2, by contrast, lacks colocalisation, gains comparator support only through PLEIO and is nominally associated across three FinnGen endpoints in two clinical families. Of the ten loci with evidence in all three streams (Results), DALRD3 reached q<0.001 without any single-trait association meeting corrected gene-level significance, with its distributed profile providing cross-trait resolution within the established 3p21.31 RHOA-region coronary-artery-disease locus [59, 60]. Because colocalisation, FinnGen external support and prior multi-trait studies address different questions (a shared regulatory variant, independent endpoint association, and convergence with published analyses), support concentrated in one stream and absent in another is complementary rather than contradictory. Absence of a given evidence type is not evidence against a locus. Table 3 turns the aggregate evidence into locus-level priorities for follow-up; statistical representatives are locus labels, not causal-gene assignments.

Beyond the stringent core, several loci stood out for their distributed profiles. The clearest is MMP16 (CTAS=18.00, q=0.0029), which had the highest participation ratio of any locus (9.41) and was retained in all 16 leave-one-trait-out analyses, yet no constituent trait reached MAGMA P<0.001 and only coronary artery disease reached P<0.01. Its signal is distributed across decorrelated dimensions and is coherent with vascular-smooth-muscle extracellular-matrix remodelling, given reported membrane-type matrix metalloproteinase expression in vascular smooth muscle cells, and MMP16 has been identified, with replication, as a coronary-artery-calcification locus [68, 69]. This offers a plausible vessel-wall hypothesis for follow-up. The ZNF471 region (CTAS=18.45, q=0.0019, participation ratio 7.26; 16/16 leave-one-trait-out) is an emerging locus: moderate colocalisation for ZNF471 in atrial appendage with cardiomyopathy (PP4=0.75), whereas FinnGen support is limited (no locus member reached P<0.01 in more than one endpoint), and the effector among ZFP28, ZNF470 and ZNF471 is unresolved. At the EFR3B-labelled locus, ADCY3 and DNAJC27 were also CTAS-significant, and ADCY3 is the clearest established cardiometabolic candidate [70]. The locus had a high participation ratio (PR=8.17) and remained significant in all 16 leave-one-trait-out analyses despite lacking a dominant single-trait signal. In FinnGen, its members were associated (P<0.01) with AF and hypertension (EFR3B) and with angina, coronary atherosclerosis and hypertension (ADCY3, which met the three-endpoint criterion). CTAS prioritises the distributed locus profile but does not resolve which member is causal.

Several limitations define the scope of interpretation. The transformed rank-correlation matrix captures pairwise dependence under a Gaussian working model and may not represent all higher-order shared structure [27]. ZCA-cor whitening, retention of positive residual evidence and the magnitude–breadth product are modelling choices, although calibration and sensitivity analyses support the resulting statistic. Because prioritisation is conditional on the prespecified trait panel, coherent signals confined to highly correlated trait clusters may be down-weighted, and different panel compositions may prioritise different loci. MAGMA assigns variant evidence to gene bodies [26], and the use of one statistical representative per locus provides reproducible units without identifying causal genes, variants or mechanisms. Colocalisation was evaluated only for the statistical representative of each locus, so alternative member genes were not systematically tested. It was also limited to seven GTEx datasets, while literature comparisons used heterogeneous phenotypes and matching units. The FinnGen analysis represented six of the eight CTAS clinical families; valve disease and aortic/peripheral vascular disease were not represented, and phenotype definitions were not identical between cohorts. Finally, discovery used European-dominated Pan-UK Biobank meta-analysis P values with a European linkage-disequilibrium reference, while colocalisation used European-ancestry effect estimates. These ancestry choices constrain generalisability and motivate evaluation across more diverse cohorts [24, 71]. This study applies CTAS to one 16-trait cardiovascular panel; other phenotype domains, ancestries and gene-mapping strategies will test its scope.

A further consideration concerns power heterogeneity across the input GWAS. The Pan-UK Biobank cardiovascular endpoints differ substantially in effective sample size, and the cerebrovascular and venous-thrombotic phenotypes are less well powered than the major coronary endpoints. Domains carried chiefly by the less-powered traits may be underrepresented in the observed signal even though every trait was retained in the panel; larger or better-powered GWAS in these domains could reveal additional cross-domain structure.

These results position CTAS as a framework for cardiovascular target prioritisation. The prioritised, dependence-adjusted loci show shared cardiovascular relevance and nominate candidate targets with potential relevance across more than one cardiovascular indication, and identify specific loci for molecular and experimental resolution. Each locus can be taken forward by integrating CTAS with colocalisation, fine-mapping, molecular-QTL, perturbation and functional-screen evidence to move from a prioritised region towards a causal gene and mechanism. The stringent, multi-family loci with cross-endpoint support generate testable biomarker hypotheses for cardiovascular multimorbidity and stratified prevention. The accompanying interactive Explorer exposes the locked gene- and locus-level results with their family profiles and support layers, enabling systematic, reproducible target-discovery workflows across the full result. CTAS indicates where this work should begin; causal genes, mechanisms and therapeutic potential remain to be established.

## Conclusions

CTAS offers a general way to extract calibrated gene-level evidence from correlated trait panels without discarding clinically meaningful structure. In cardiovascular disease, it turned 16 correlated traits into FDR-controlled gene and locus sets that were stable, increasingly multi-family and supported by molecular data, previous studies and FinnGen. These loci provide focused priorities for fine-mapping, molecular-QTL integration and experimental perturbation [5, 7, 72]. More broadly, it provides a practical bridge from correlated disease-association data to the functional work that turns shared genetic signals into biology and targets.

## Declarations

## Ethics approval and consent to participate

This study analysed only publicly available, summary-level genetic association statistics (Pan-UK Biobank, FinnGen release 12) and publicly available expression quantitative trait locus summary statistics (GTEx v8 via the eQTL Catalogue). No individual-level data were accessed. The contributing studies obtained ethical approval and informed consent as described in their original publications. No additional ethical approval was required for this secondary analysis of summary statistics.

## Consent for publication

Not applicable.

## Availability of data and materials

The datasets supporting the conclusions of this article are included within the article and its supplementary files (Supplementary files 1–13). The analysis code, derived inputs, results and interactive Explorer are available in the GitHub repository https://github.com/richard-monaghan/ctas-cardiovascular, archived at Zenodo https://doi.org/10.5281/zenodo.24499456. The interactive Explorer is available at https://richard-monaghan.github.io/ctas-cardiovascular/.

The publicly available data analysed were: Pan-UK Biobank summary statistics (https://pan.ukbb.broadinstitute.org; CC BY 4.0); FinnGen release 12 summary statistics (https://www.finngen.fi/en/access_results); GTEx v8 cis-eQTL summary statistics from the eQTL Catalogue (https://www.ebi.ac.uk/eqtl/, study QTS000015; CC BY 4.0); the 1000 Genomes Project phase 3 European reference panel; and HGNC gene nomenclature (https://www.genenames.org). The comparator studies’ source tables are available from the original publications. Raw GWAS summary statistics are not redistributed.

Software. Project name: CTAS (Cross-Trait Aggregation Score). Project home page: https://github.com/richard-monaghan/ctas-cardiovascular. Archived version: https://doi.org/10.5281/zenodo.24499456 (v1.0.0). Operating system(s): macOS or Linux. Programming language: R (≥4.5), Python (≥3.13). Other requirements: R packages Matrix, digest and cluster (coloc and data.table for colocalisation); Python packages pandas, numpy, scipy and openpyxl; MAGMA v1.10 and tabix for upstream steps. License: MIT (code); CC BY 4.0 (derived data and documentation). Any restrictions to use by non-academics: none.

## Competing interests

The authors declare that they have no competing interests.

## Funding

This work was supported by British Heart Foundation Programme Grant RG/F/21/110050. B.D.K. was supported by a British Heart Foundation personal chair. H.G. was supported by the Engineering and Physical Sciences Research Council (EPSRC) through the Causality in Healthcare AI Hub (CHAI), grant number EP/Y028856/1.

## Authors’ contributions

RMM conceived the study and developed the methodology; wrote the software and led the analyses; carried out validation; curated the data; produced the visualisations; wrote the original draft and revised the manuscript. HG contributed to the methodology through statistical review and contributed to validation; reviewed and edited the manuscript. SGW contributed to validation; reviewed and edited the manuscript. BDK supervised the work; provided resources and acquired funding; reviewed and edited the manuscript. All authors read and approved the final manuscript.

CRediT taxonomy: R.M.M.: Conceptualization; Methodology; Software; Formal analysis; Investigation; Validation; Data curation; Visualization; Writing – original draft; Writing – review C editing. H.G.: Methodology (statistical review); Validation; Writing – review C editing. S.G.W.: Validation; Writing – review C editing. B.D.K.: Supervision; Resources; Funding acquisition; Writing – review C editing.

## Acknowledgements

We thank Korbinian Strimmer for helpful discussion of the whitening approach.

We acknowledge the Pan-UK Biobank (Pan-UKB) project and UK Biobank. The Pan-UKB summary statistics were generated using the UK Biobank Resource (project ID 31063).

We want to acknowledge the participants and investigators of the FinnGen study.

The Genotype-Tissue Expression (GTEx) Project was supported by the Common Fund of the Office of the Director of the National Institutes of Health, and by NCI, NHGRI, NHLBI, NIDA, NIMH, and NINDS. The data used for the analyses described in this manuscript were obtained from the eQTL Catalogue (GTEx v8, study QTS000015) on 7 August 2026 and 16 November 2026.

We thank the eQTL Catalogue, the 1000 Genomes Project and the HUGO Gene Nomenclature Committee for making their resources publicly available.

## Notes

### Competing Interest Statement

The authors have declared no competing interest.

