## Supplementary file 1 (Supplementary Figures 1-5) for "Dependence-aware aggregation maps shared genetic architecture across cardiovascular traits"

### a Empirical P against the $10^9$-draw null

### b Benjamini–Hochberg step-up

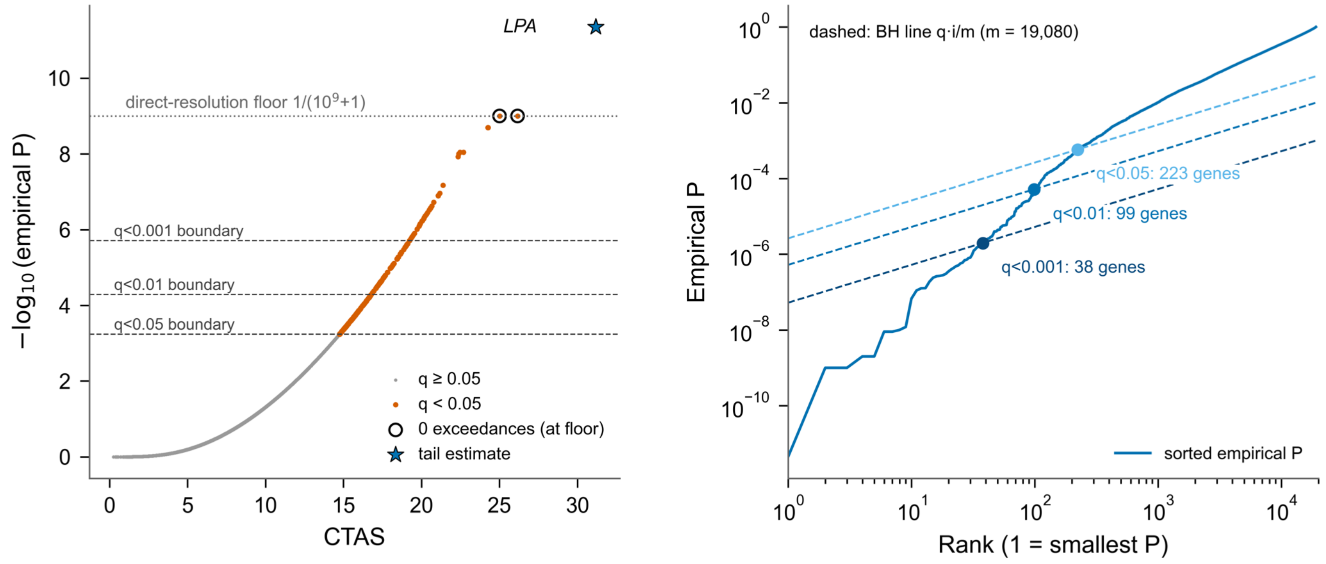


### c Most extreme genes and tail treatment

| Gene | CTAS | Null exceedances (of $10^9$) | Empirical P | P basis | BH q | q-set |
|---|---|---|---|---|---|---|
| *LPA* | 31.16 | 0 | 4.47e-12 | tail estimate | 8.52e-08 | q<0.001 |
| *CDKN2B* | 26.16 | 0 | 1e-09 | floor 1/(M+1) | 6.36e-06 | q<0.001 |
| *IGF2R* | 25.01 | 0 | 1e-09 | floor 1/(M+1) | 6.36e-06 | q<0.001 |
| *FES* | 24.27 | 1 | 2e-09 | direct count | 7.63e-06 | q<0.001 |
| *APOE* | 24.26 | 1 | 2e-09 | direct count | 7.63e-06 | q<0.001 |
| *SLC22A1* | 22.68 | 8 | 9e-09 | direct count | 2.38e-05 | q<0.001 |
| *MARK3* | 22.46 | 8 | 9e-09 | direct count | 2.38e-05 | q<0.001 |
| *SLC22A3* | 22.38 | 9 | 1e-08 | direct count | 2.38e-05 | q<0.001 |

3 genes had zero exceedances; 1 (LPA) received the tail estimate. The q-set boundaries lie at empirical P $5.8\times10^{-4}$, $5.1\times10^{-5}$ and $2.0\times10^{-6}$, far above the direct-resolution floor $1/(10^9 + 1)$, so the tail treatment changed no q-set boundary.

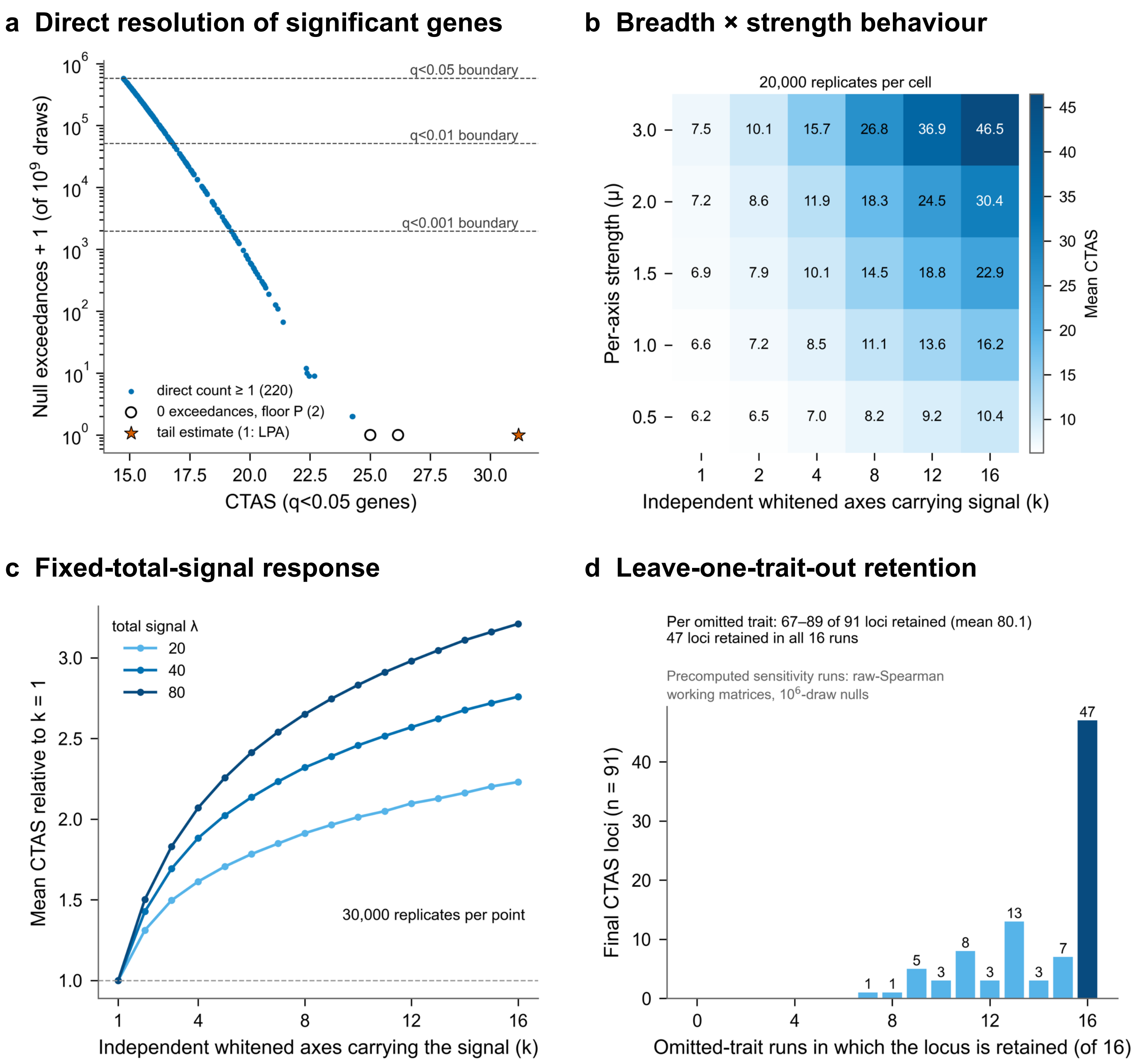

a Direct resolution of significant genes
Null exceedances + 1 (of $10^9$ draws)
$10^6$
$10^5$
$10^4$
$10^3$
$10^2$
$10^1$
$10^0$
q<0.05 boundary
q<0.01 boundary
q<0.001 boundary
direct count ≥ 1 (220)
0 exceedances, floor P (2)
tail estimate (1: LPA)
15.0
17.5
20.0
22.5
25.0
27.5
30.0
CTAS (q<0.05 genes)
b Breadth × strength behaviour
20,000 replicates per cell
Per-axis strength (μ)
3.0 7.5 10.1 15.7 26.8 36.9 46.5
2.0 7.2 8.6 11.9 18.3 24.5 30.4
1.5 6.9 7.9 10.1 14.5 18.8 22.9
1.0 6.6 7.2 8.5 11.1 13.6 16.2
0.5 6.2 6.5 7.0 8.2 9.2 10.4
1 2 4 8 12 16
Independent whitened axes carrying signal (k)
Mean CTAS
45
40
35
30
25
20
15
10
c Fixed-total-signal response
total signal λ
20
40
80
Mean CTAS relative to k = 1
3.0
2.5
2.0
1.5
1.0
30,000 replicates per point
1
4
8
12
16
Independent whitened axes carrying the signal (k)
d Leave-one-trait-out retention
Per omitted trait: 67–89 of 91 loci retained (mean 80.1)
47 loci retained in all 16 runs
Precomputed sensitivity runs: raw-Spearman
working matrices, $10^6$-draw nulls
Final CTAS loci (n = 91)
40
30
20
10
0
1 1 5 3 8 3 13 3 7 47
0
4
8
12
16
Omitted-trait runs in which the locus is retained (of 16)

### a  Signed whitened profiles of the 91 locus representatives

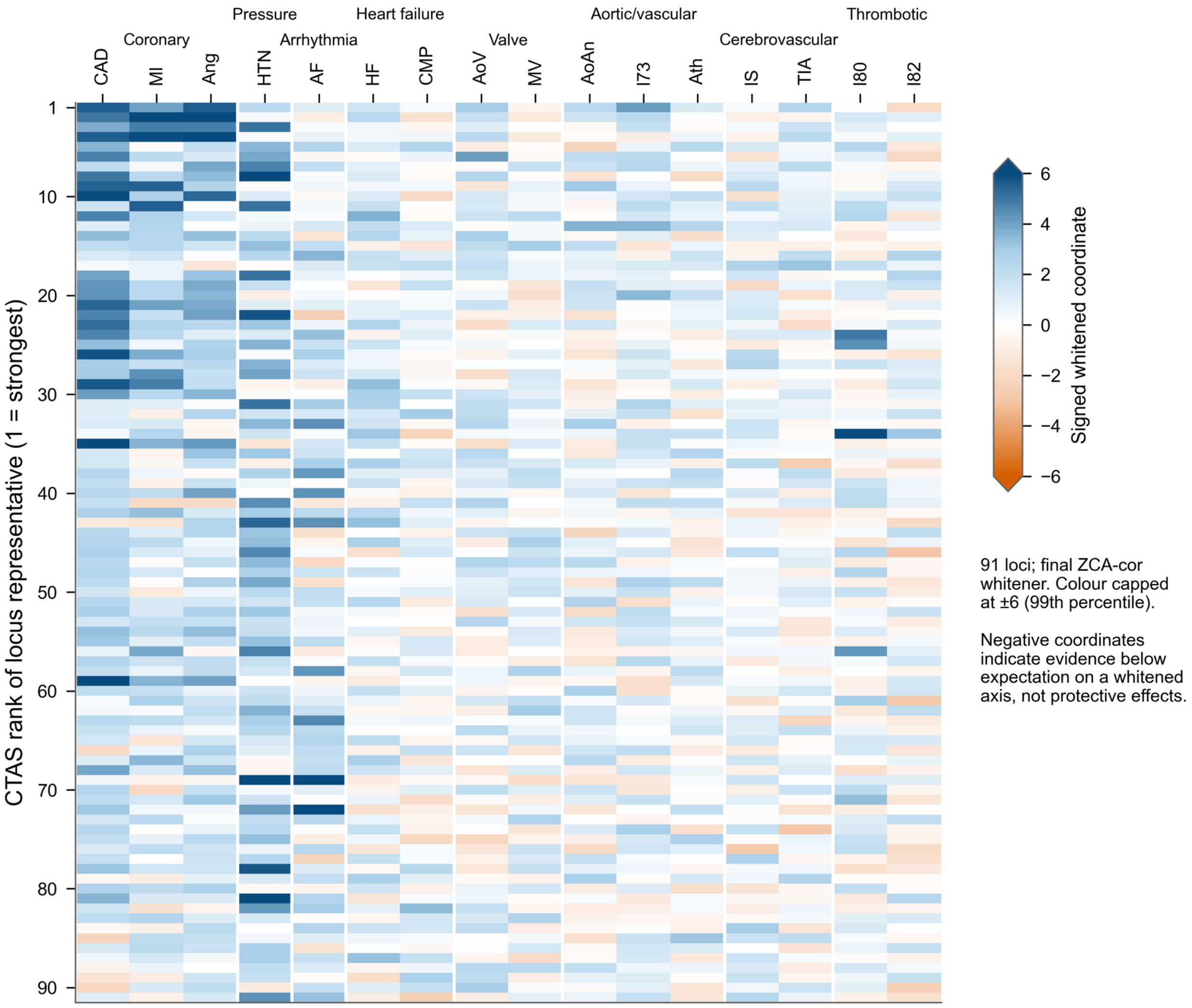


### b  Ordination of family composition

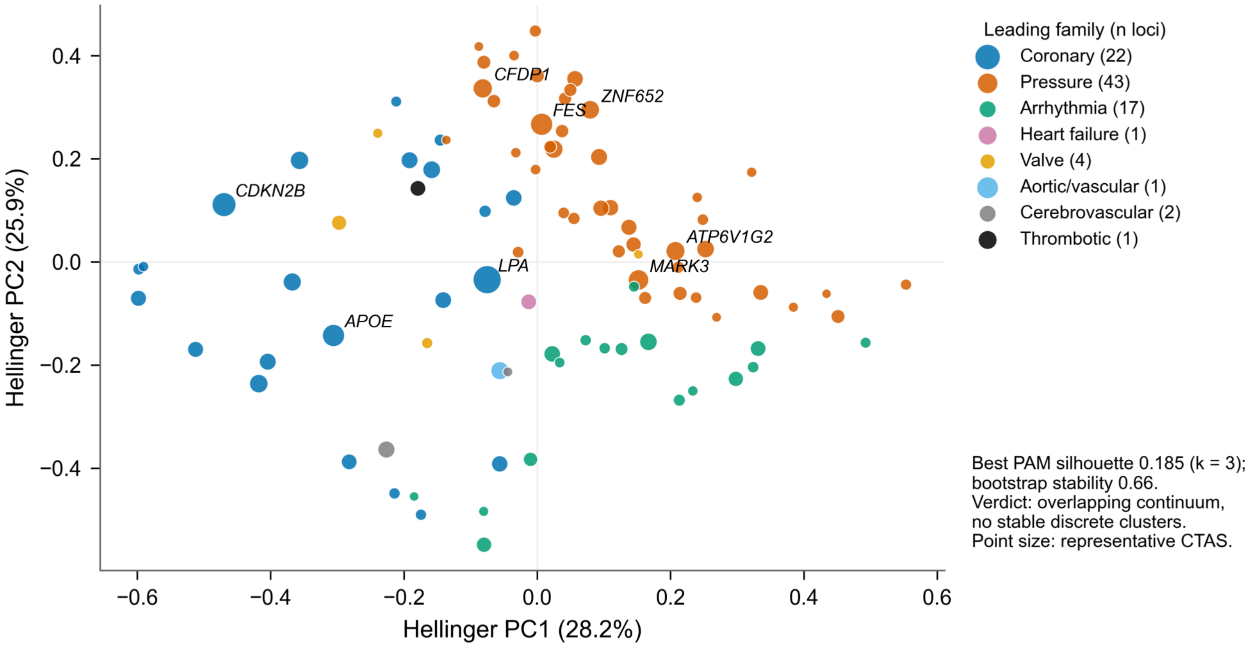

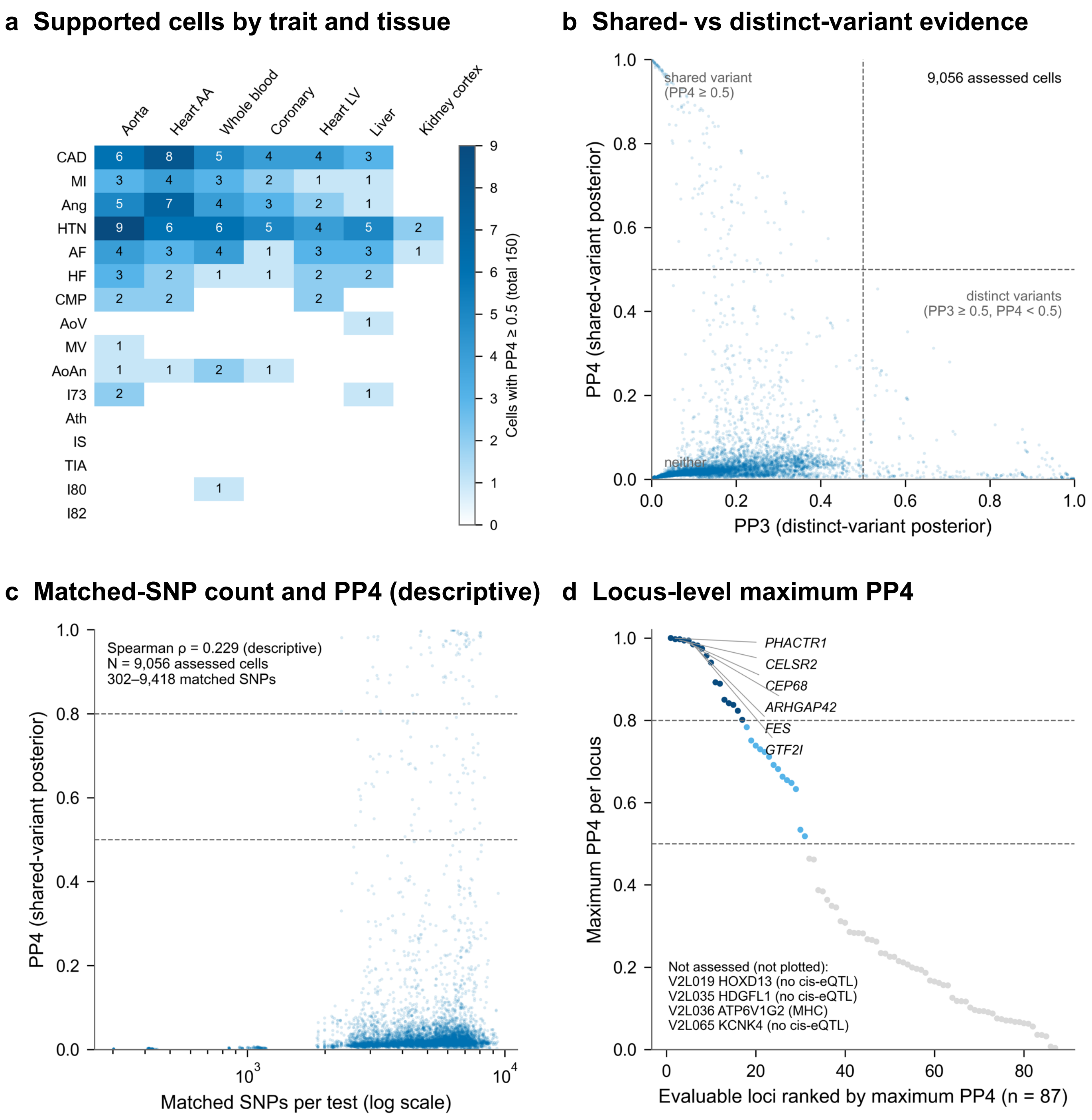
a Supported cells by trait and tissue
Aorta
Heart AA
Whole blood
Coronary
Heart LV
Liver
Kidney cortex
CAD
MI
Ang
HTN
AF
HF
CMP
AoV
MV
AoAn
I73
Ath
IS
TIA
I80
I82
Cells with PP4 ≥ 0.5 (total 150)
b Shared- vs distinct-variant evidence
shared variant
(PP4 ≥ 0.5)
9,056 assessed cells
distinct variants
(PP3 ≥ 0.5, PP4 < 0.5)
neither
PP4 (shared-variant posterior)
PP3 (distinct-variant posterior)
c Matched-SNP count and PP4 (descriptive)
Spearman ρ = 0.229 (descriptive)
N = 9,056 assessed cells
302–9,418 matched SNPs
PP4 (shared-variant posterior)
Matched SNPs per test (log scale)
d Locus-level maximum PP4
PHACTR1
CELSR2
CEP68
ARHGAP42
FES
GTF2I
Not assessed (not plotted):
V2L019 HOXD13 (no cis-eQTL)
V2L035 HDGFL1 (no cis-eQTL)
V2L036 ATP6V1G2 (MHC)
V2L065 KCNK4 (no cis-eQTL)
Maximum PP4 per locus
Evaluable loci ranked by maximum PP4 (n = 87)

### a Four-comparator intersections across the 91 CTAS loci

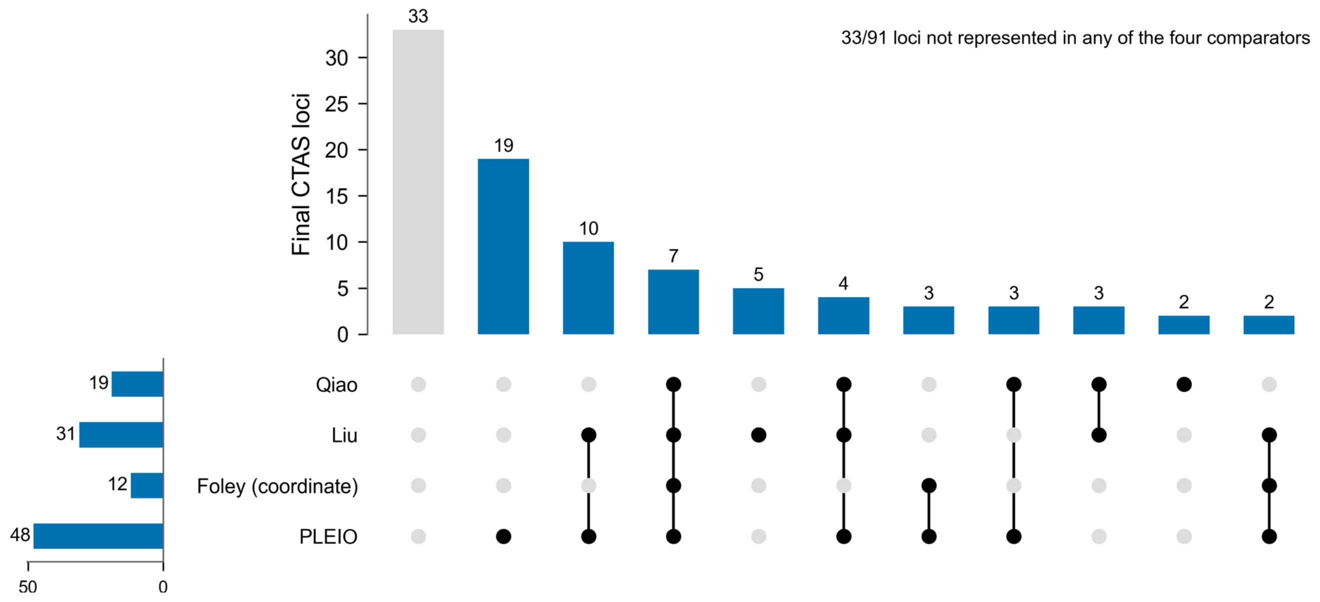


### b Gene-level FinnGen support by threshold

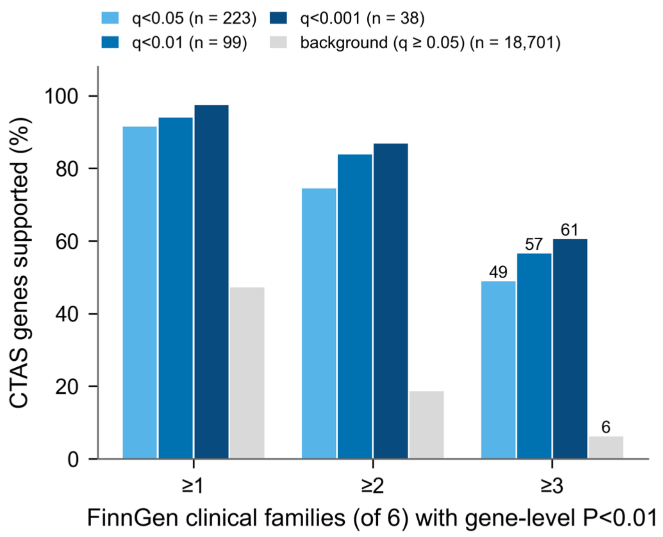


### c FinnGen family counts, q<0.05 genes

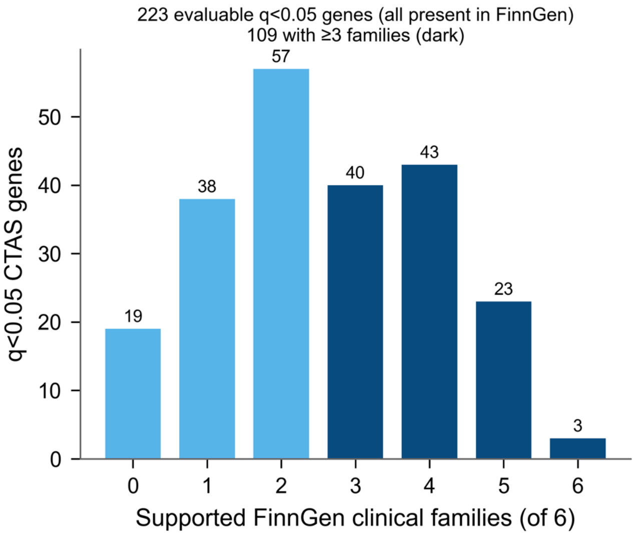
